# Measuring the impact of nonpharmaceutical interventions on the SARS-CoV-2 pandemic at a city level: An agent-based computational modeling study of the City of Natal

**DOI:** 10.1101/2022.05.05.22274749

**Authors:** Paulo Henrique Lopes, Liam Wellacott, Leandro de Almeida, Lourdes Milagros Mendoza Villavicencio, André Luiz de Lucena Moreira, Rislene Katia Ramos de Sousa, Priscila de Souza Silva, Luciana Lima, Michael Lones, José-Dias do Nascimento, Patricia A. Vargas, Renan Cipriano Moioli, Wilfredo Blanco Figuerola, César Rennó-Costa

## Abstract

The severe acute respiratory syndrome coronavirus 2 (SARS-CoV-2) pandemic hit almost all cities in Brazil in early 2020 and lasted for several months. Despite the effort of local state and municipal governments, an inhomogeneous nationwide response resulted in a death toll amongst the highest recorded globally. To evaluate the impact of the nonpharmaceutical governmental interventions applied by different cities – such as the closure of schools and business in general – in the evolution and epidemic spread of SARS-CoV-2, we constructed a full-sized agent-based epidemiological model adjusted to the singularities of particular cities. The model incorporates detailed demographic information, mobility networks segregated by economic segments, and restricting bills enacted during the pandemic period. As a case study, we analyzed the early response of the City of Natal – a midsized state capital – to the pandemic. Although our results indicate that the governmental response could be improved, the restrictive mobility acts saved many lives. The simulations show that a detailed analysis of alternative scenarios can inform policymakers about the most relevant measures for similar pandemic surges and help developing future response protocols.

## Introduction

The severe acute respiratory syndrome coronavirus 2 (SARS-CoV-2) pandemic started in Wuhan, China, in December 2019 [1–3] and quickly spread outside its borders, being acknowledged as a worldwide event by the World Health Organization in March 2020 [4]. The associated Covid-19 disease has a case fatality ratio of around 1.5% and is substantially more deadly for the elderly [5–7], imposing significant pressure on public health systems. By the end of the year of 2021, the pandemic had claimed the life of about 18 million people worldwide [8]. Effective vaccines were only widely deployed in 2021, with a delayed distribution in underdeveloped countries [9, 10]. The transmission of SARS-CoV-2 is mainly airborne, with a relatively high estimated ratio of transmissions originated from asymptomatic or presymptomatic infected people [11–14]. Since testing availability differs significantly across countries, and because tests are only effective for a limited window of the infectious cycle, it is difficult to identify the infectious vectors. It is, thus, considerably challenging for public health operators to establish effective mitigation policies. Different countries applied a myriad of nonpharmaceutical interventions to reduce the impact of the pandemic, such as the closure of businesses and public services, the obligation of the wear of face masks, and limitations in mobility (lockdown). However, the inability to assess their effectiveness and the critical economic and social impact of such measures imposes substantial political costs to policymakers, which, in turn, often opt to drop them off based on nonscientific arguments.

Establishing an optimal response strategy to the pandemic is especially problematic in Brazil - the largest and most populated country in Latin America, with over 211 million inhabitants [15]. The virus was introduced in Brazil from Europe between February and March 2020 [16]. After an early phase with locally constrained spread, the pandemic affected all regions of Brazil, with the first peak of deaths in June 2020 followed by a slow decay that reverted in November 2020. As by the end of 2021, over 600 thousand deaths were reported in Brazil, with a peak 7-day average daily casualties rate above 3000 deaths per day in April 2021 [17]. The pandemic in Brazil had two preeminent waves marked by high contamination levels and mortality, with more robust social mobilization in the first wave and higher impact rates in the second wave [8].

It is hard to evaluate the impact of the contention measures applied at a national level due to the heterogeneity of local responses. Brazil is a federal republic divided into 27 federation units (26 states and one federal district) and 5568 cities. The Brazilian constitution assigns to the state governors and mayors the obligation to define sanitary measures in events such as a pandemic, with coordination from the Federal government through the Ministry of Health. However, political differences between the federal and local governments resulted in each state and city following an isolated agenda. Therefore, considering that the Federal Supreme Court recognised the right of each municipality to determine its own policies and given the extensive differences within single states - the State of São Paulo has a similar population size to Spain [18] -, it is more beneficial to evaluate the impact of the contention measures at a municipal level.

Epidemiological models can help predict the impact of contention measures at a municipal level, but there are limitations. A key factor is that different cities have different mobility and hospital structures. In Brazil, most cities do not have a high-density mobility system and there are separate private and public health systems. Also, likewise most Brazilian urban centres, fairly isolated wealthy neighbourhoods contrast with highly-dense, no sanitation, poor districts. Importantly, current modelling strategies highlight that such characteristics have a significant impact on model performance. For instance, one of the first models focused on the Chinese city of Wuhan outbreak, a city with more than 11 million people, large mass transportation systems, hot summers and cold winters [19]. A following modelling study focused on US temperate regions, and authors stress that the epidemics in tropical regions can be much more complex [20]. Also, the landmark model from Imperial College [21], used as reference worldwide, emphasises that their results relate to US and UK data, with possible extension to high-income countries. Therefore, epidemiological models inspired by high levels of metropolitan transportation and more social homogeneous societies might not reflect the epidemic dynamics of small and medium cities in underdeveloped countries. Considering the exponential dynamics that are common to epidemiological models, minor amendments to parameters or model architecture may incur a significant error with severe impact on the health system and economics. Moreover, for reasons mentioned above, nationwide epidemiological models present limited information for the policymakers.

Here we use computational epidemiological modelling to assess the impact of governmental nonpharmaceutical interventions at the City of Natal, Brazil. The City of Natal is the capital of the State of Rio Grande do Norte, in the northeastern region of Brazil. With a total of 890 thousand inhabitants estimated in 2020 [15], Natal is among the 20 most populous cities in Brazil, located entirely in an urban area, with a territorial extension of 167,401 km^2^, and a demographic density of 5325,8 inhabitants per km^2^ (6th largest among Brazilian capitals). The municipality stands out as an important Brazilian tourist destination due to its beautiful beaches, lagoons, and dunes, receiving around 2 million visitors from other parts of Brazil and the world annually. In economic terms, the service sector stands out in the municipality, with the Gross Domestic Product (GDP) being the 16th in the ranking of the capitals of the 27 federation units of the country [18]. In terms of well-being, Natal was in 2010 among the 100 Brazilian municipalities with the highest Human Development Index (HDI) (0.763) [22]. However, the municipality has important social vulnerabilities that seem to have been amplified with the sanitary emergency, as the unemployment rate reached 13.8% in the first three months of the pandemic [23] while the most rigid social distancing prevailed. We have chosen the City of Natal due to the availability of well-documented epidemiological and geographical data. It shares similarities with many other Brazilian cities, such as population size, urban organisation, and policies undertaken during the pandemic.

In contrast to the commonly used compartmental models [24–27], we built an agent-based model that allows the inclusion of detailed demographic information that is commonly available, enabling easy replication of the analysis for other cities. Furthermore, agent-based models are superior in capturing complex heterogeneous urban and social interactions during infection outbreak [28, 29]. The model represents social interaction probabilities as graphs that display similar properties of the census data. Such a complex network approach allows segmenting the modelled interaction between age groups and social modalities, such as religion, work, and school, to assess the impact of individual measures. In our strategy, we establish a baseline model that reproduce the observed case fatality curve. Next, we modify model parameters to emulate alternative scenarios in which different actions were taken by the administration and evaluate the attainable fatalities outcome. Through this strategy, we can quantify the impact – in terms of lives saved or lost – if different policies had been applied.

Our results indicate that the policies enforced by local government could be significantly improved but nevertheless prevented a much more catastrophic scenario. While early decisions to close schools and universities saved many lives, reopening commerce and religious gatherings came with a substantial cost of lives.

## Materials and Methods

### Epidemiological and demographic data

Epidemiological data of the pandemic in the State of Rio Grande do Norte (RN) has been made available online by the State Secretary of Health (SESAP-RN). The daily bulletins report the anonymous data as age and gender of all confirmed cases, tests, hospitalisations, and deaths. For this study, we considered only the first epidemiological wave, defined here as two weeks before the first reported case on March 12th 2020 to November 4th 2020 when the average 7-day death rate was below 1 case per day for the first time. Data of hospitalization and ICU beds occupancy are also available from a different source of SESAP-RN (Regula-RN system). No information was available about the place of residence of the ICU occupants.

The total number of Natal city residents and age distribution was based on the urban plan from 2017 [30]. Demographic data were collected from official governmental sources, and these were used to describe the main social activities that were affected by the social distancing decrees during the pandemic, such as: home, work, transport, religion, basic and higher education. To describe the interactions within the household, we used the total number of family members calculated from the Continuous National Household Sample Survey - Continuous PNAD, a survey of national coverage and representative for Brazilian capitals and states [23]. Considering the workplace as an important space for disseminating the virus and directly affected by the decrees of social distancing, we used data from the Annual List of Social Information – RAIS [31]. In Brazil, employers provide the administrative records of workers in the formal sector to the Ministry of Economy. In this article, five economic sectors were considered: agricultural, industrial, construction, commerce, and service [31]. To describe the circulation activities, information from the last demographic census was taken into account to obtain the average time spent by residents of the municipality for work [32].

Furthermore, information related to the carrying capacity of public transport in the City of Natal, provided by the Municipal Department of Urban Mobility [30], was used. The 2010 Demographic Census was also used to obtain the proportion of people who declared themselves Catholics and Evangelicals. These two religious categories were considered because they are, respectively, the most predominant in the Brazilian population [32]. Concerning educational establishments, Brazil has two sources of official census information: (1) the Brazilian School Census, from which the total number of students enrolled from kindergarten to professional education in public and private education in the City of Natal [33]; and (2) the Higher Education Census, from which we obtained information on the total number of students at the Federal University of Rio Grande do Norte on the Natal campus [34].

Notably, according to Brazilian Law, the study does not require an Ethical Committee evaluation as all data used is anonymized and is in the public domain.

### Epidemic agent-based model

We developed an agent model that extends the classic SIR model [35] with a more specific list of health states and with an agent-level individual interaction mechanism that implements social layers as complex networks [36]. This algorithm has two core parts into simulation: (1) the infection model implements a state machine that tracks the health of each agent across the simulation with respect to the expected disease course; and (2) a complex interaction network that simulates the agent interactions within multiple social layers (e.g. contact at work, transport, schools, etc.). In addition to that, we simulate the City of Natal population (a total of 873383 agents) over 253 days (February 26th to November 4th of 2020), the first wave of the pandemic. This section summarises the simulation implementation. The simulation code, data, and user documentation can be found at Github.

#### Agent dynamics

In order to better emulate the Covid-19 infection cycle, we extended the classic SIR model with additional states:

- **S**usceptible – The agent is vulnerable to Covid-19 and may be infected any day. Initially all agents are in this state.
- **I**ncubated – The agent has been exposed to the disease however is not yet exhibiting symptoms.
- **A**symptomatic - The agent has the disease past the incubation period, however does not show any symptoms.
- **I**mmune – The agent was infected and recovered; In our model we assume once an agent has been infected, they develop immunity and cannot be reinfected. The agent may also have a natural immunity to the disease.
- **S**ymptomatic Mild – The agent was infected and has mild symptoms such as cough and sore throat.
- **S**ymptomatic Moderate – The agent was infected and has moderate symptoms. In addition to cough and sore throat (but not exclusively), the agent had headache and fever. They do not need hospitalisation at this stage.
- **H**ospital – The agent was infected, the symptoms continued to develop and they must be hospitalised.
- **I**CU – The agent is already hospitalised in critical condition and needs ICU and ventilation support.
- **D**ead – The agent developed severe symptoms and did not survive.

Initially, all agents are in a *Susceptible* health state. Agents get infected by close contact with another infectious agent by following the interaction rules of the complex networks (see below). Once infected, the health state of the agent turns into *Incubated*, starting a sequence of transitions through the different health states which will ultimately end as an *Immune* individual or a new *Death* (Fig 1A). For each infected agent, the model randomly assigns the most serious health state the agent will reach – and therefore also whether the agent will eventually die or not – following the probabilities that are presented in Table (1). The outcome likelihood is modulated by the age of the agent as reported by the State Secretary of Public Health (SESAP-RN) and were computed as the proportion of the number of confirmed cases by health state (*Symptomatic Mild, Symptomatic Moderate, Hospital, ICU* and *Dead*); for each age range. The odds for *Asymptomatic* cases were based on the proportion proposed by [37], resulting on a value of 18.8% that was applied on all confirmed cases.

**Fig 1.**
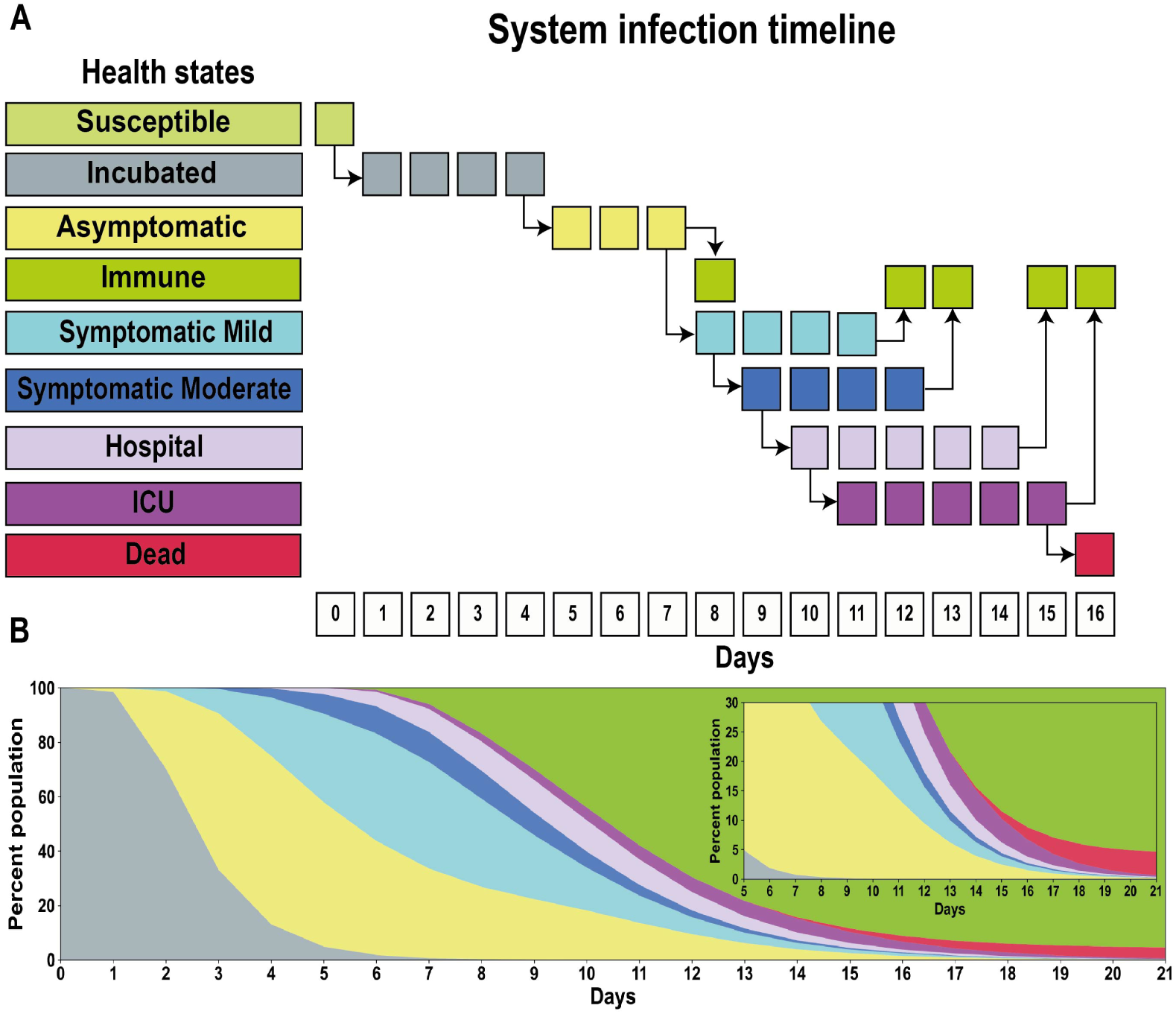
Health states timeline and proportion of the Covid-19 implemented on the system. On average, 16 days are necessary for an agent to reach the *Dead* health state. Once they develop the disease as *Incubated*, the agent remains on average four days before changing to the *Asymptomatic* health state (time average is three days at this state). After this state, the agents may become *Immune* or continue to evolve the disease and reach the *Symptomatic Mild* health state, where four days pass before changing the state to *Immune*. Whether the disease continues to evolve, the agent already turns to the health state to *Symptomatic Moderate* on the first day. To become *Immune*, the agents have to pass four days at *Symptomatic Moderate*. Whether the pathology evolves (already on the first day), the agent needs to go to the *Hospital*, where pass five days to reach the *Immune* health state. The agent dead occurs only after five days at *ICU* treatment. The number of ICU beds available on the system is unlimited. The B panel represents the spread of disease among agents (a total of 10000). Initially, all agents begin at Incubated health state and over the days, the pathology evolves as shown in A panel.

**Table 1.** Health state probability disease by age range. Each health state is related to an age range, showing the outcome state that an agent reaches in case of infection.

|  |  | Age range |  |  |  |  |  |  |  |  |
| --- | --- | --- | --- | --- | --- | --- | --- | --- | --- | --- |
|  |  | 0-9 | 10-19 | 20-29 | 30-39 | 40-49 | 50-59 | 60-69 | 70-79 | 80+ |
| Final stage of Health | Asymptomatic | 0.1579 | 0.1587 | 0.158 | 0.1583 | 0.1582 | 0.1584 | 0.1583 | 0.1583 | 0.1583 |
|  | Symptomatic Mild | 0.4269 | 0.5904 | 0.6226 | 0.6088 | 0.5647 | 0.4421 | 0.3442 | 0.1883 | 0.106 |
|  | Symptomatic Moderate | 0.0819 | 0.1513 | 0.1682 | 0.1348 | 0.1041 | 0.0854 | 0.0536 | 0.0215 | 0.0102 |
|  | hospital | 0.3333 | 0.0996 | 0.0512 | 0.981 | 0.173 | 0.3142 | 0.4439 | 0.6319 | 0.7255 |
|  | ICU | 0.0643 | 0.0185 | 0.0096 | 0.18 | 0.0412 | 0.0961 | 0.1603 | 0.2847 | 0.3355 |
|  | Dead | 0.0234 | 0.111 | 0.0084 | 0.0135 | 0.0292 | 0.0742 | 0.1147 | 0.2221 | 0.2752 |

The time an agent remains in the *Incubated* state is stochastic and follows a log-normal distribution of the agents’ average disease incubation time [38]. The length of all other transitions is deterministic as they do not affect transmission dynamics but are defined based on the individual incubation time. As a result, a different proportion of health states develop in the population following the initial day of infection (Fig 1). On average, the agents spent four days at *Incubated* state before evolving to *Asymptomatic*, where it remains for three days. Once at *Asymptomatic* health state, the agent may be able to become *Immune* or develop symptoms and reach the *Symptomatic Mild* health state, wherein it has to pass four days before becoming *Immune* [39]. *However, already on the first day at this health state, the symptoms could develop and the health state changes to Symptomatic Moderate*, in which the agent has to spend four days [40] before becoming *Immune*. If the symptoms become serious, the agent pursues *Hospital* health assistance. After one day at the hospital, the agent may be in critical conditions and require ICU support or ventilation; these require five days of intensive care. The time duration in hospitalisation and ICU was based on the State Secretary of Public Health (SESAP-RN). Importantly, the number of hospital or ICU admissions on the model is unlimited, i.e., all agents can be admitted at once if necessary.

Each agent is contagious in a specific infectious period that starts four days after the contamination (on average already at the *Incubated* health state) and finish two days before the final incubation time in the *Asymptomatic* health state. If the agent progresses to *Symptomatic Mild*, it remains infectious up to two days before the end of this health state. These three health states are the only in which the agent can transmit the disease, which is an optimistic assumption because it considers that 1) once the agent reaches the *Symptomatic Moderate* it will be isolated, and 2) that the agents that progress to *Hospital* will not infect the health staff.

#### Complex Networks of Social Interactions

At each cycle of the simulation (day), the system computes the possible social interactions between agents of the population to emulate the contagious aspect of the pandemic. As a result of this interaction, one agent in the infectious period can infect another susceptible agent and change its health state into incubated. The probabilities of interaction are determined by multiple complex networks of social interactions (layers and sub-layers) that include an agglomerate of agents. Initially, when the agents are created, they are distributed to the layers/sub-layers according to the age range (Table 2, third column) and the probability of belongingness (Table 2, fourth column). The time per week during which social interactions happen in each layer/sub-layer are displayed on the fifth column. Once allocated, they are separated into sub-groups to represent the many activities clusters. The average direct contact among agents and the group size is presented in columns six and seven, respectively. Each network has its specificities and is further detailed in Table (2). The probability of agents interacting among themselves (referred throughout the text as *P*_*interaction*_) within layers/sub-layers is calculated over time as follows. Consider two agents *A*_1_ and *A*_2_ belonging to the same sub-layer, its interaction value is calculated by:

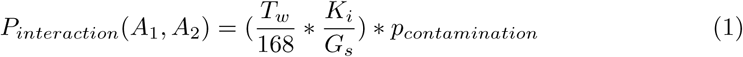

**Table 2.** Social interaction layers information. The first column (Layer) shows the layers (total of 7) implemented on the system, where some needed sub-layers (schools, work and religion) to represent the activities better (second column). The third column (Agents age) represents the age range distribution for each layer/sub-layer. The fourth column represents the probability of agents belong to the layer. In some sub-layers (schools and work), the sum of this probability is 100%, which means that all agents with age range necessary belong to only one of these sub-layers. The fifth column (Time per week) is the average time the agents spend interacting with other agents on this layer/sub-layer. The sixth column (Nearest) shows the agent’s average number of contacts per layer/sub-layer. The last column (Group size) represents the average groups’ size and the agent’s distribution; per layer/sub-layer.

| Layers | Sub-layers | Agents age | Belongingness probability [%] | Time per week ($T_w$) | Nearest ($K_i$) | Group size ($G_s$) |
| --- | --- | --- | --- | --- | --- | --- |
| Schools | Kindergarten public | 0-9 years | 54.53 | 20 hours | 8 | [10-55] Norm |
|  | Kindergarten private | 0-9 years | 45.47 | 20 hours | 2 | [3-24] Norm |
|  | Elementary public | 10-19 years | 62.36 | 20 hours | 7 | [8-32] Norm |
|  | Elementary private | 10-19 years | 37.64 | 20 hours | 4 | [11-19] Norm |
|  | Professional public | 20-29 years | 56.43 | 25 hours | 5 | [6-64] Log-norm |
|  | Professional private | 20-29 years | 43.57 | 25 hours | 4 | [6-39] Norm |
| Work | Agriculture | 20 - 69 years | 0.87 | 40 hours | 5 | [6-41] Norm |
|  | Industry | 20 - 69 years | 11.96 | 40 hours | 5 | [6-37] Norm |
|  | Construction | 20 - 69 years | 7.12 | 40 hours | 5 | [6-38] Norm |
|  | Commerce | 20- 69 years | 22.91 | 40 hours | 4 | [5-35] Norm |
|  | Services | 20- 69 years | 57.13 | 40 hours | 5 | [6-37] Norm |
| Home | Home | Everyone | Everyone | 21 hours | Everyone | [2-13] |
| Transport | Transport | 10 - 80+ years | 31 | 1h44min | 2 | [3-70] Uniform |
| Religion | Catholic | 0 - 69 years | 54.77 | 2 hours | 8 | [17-100] Norm |
|  | Evangelic | 0 - 69 years | 17.08 | 2 hours | 9 | [18-100] Norm |
| UFRN | UFRN | 10 - 69 years | 4.1 | 40 hours | 5 | [12-180] Norm |
| Random | Random | Everyone | 5 per person | 1 hour | 1 | 1-to-1 |

Where *T*_*w*_ is the time per week spent by the agents into the sub-layer, *K*_*i*_ is the average of direct contacts with other agents, 168 is the total hours in one week (24 hours * 7 days = 168 hours) and *G*_*s*_ is the group size inside the sub-layer. The *P*_*contamination*_ value represents the probability of infection upon contact, i.e., the probability of virus spread. Initially, this value is 1.7 and was obtained from (SESAP-RN) as the average of *R*_0_ value for the first wave. Each layer/sub-layer has its own *P*_*contamination*_ and this value is modified over the days as consequence of the decrees (more details are presented in Decrees sub-section).

These complex networks of social interactions were built based on demographic data detailed in Table (2). The structural differences among the different layers and sub-layers become evident by plotting the agents and their social interactions (Fig 2).

**Fig 2.**
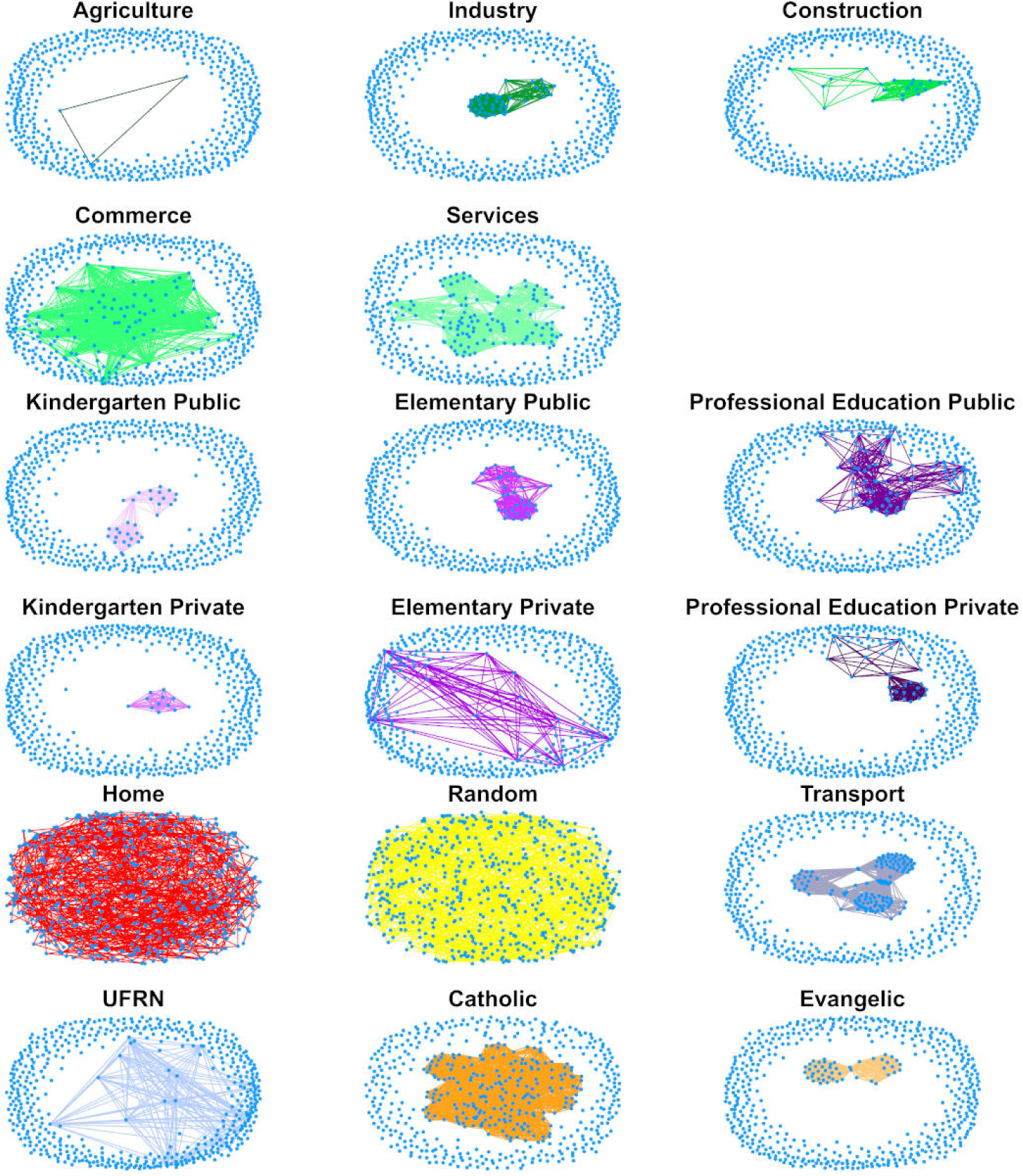
Complex network layers and connections. Each plot shows one layer/sub-layer (total of 17). The plots’ blue dots represent the agents (total of 550) and the lines indicate their connections (social interaction). The layers Home and Random are the most connected once all agents have connections in this layer (minimum of 1 for Home layer and 5 for Random); agriculture has the lowest number of connections. All the information about connections is available in Table (2).

To better represent the social interaction of the City of Natal population, we divided some activities into sub-layers, such as Work, School and Religion (Table 2, second column). The first six layers (Agriculture, Industry, Construction, Commerce and Services) belong to the “Work layers” group. The following six layers (Kindergarten, Elementary and Professional Education; public and private) correspond to “Schools layers”. The last five layers (Home, Random, Transport, UFRN, Catholic and Evangelic) belong to the “Other layers” group.

The School layer was split into six sub-layers to represent the educational system in Natal. This layer is composed of: public and private kindergarten, elementary, and professional education. Each agent may only be a part of one sub-layer according to their age range (Table 2, the third column). The fourth column shows the probability of an agent belonging to each sub-layer. These probabilities add to 100% in each educational age range for public and private. All groups follow a normal distribution, except the Professional public education layer, where the individuals in this layer have a log-normal distribution. The data used in this layer comes from 2019 Brazilian School Census [33].

To represent the Work layer, formal employment in five economic sectors were considered: agricultural, industrial, construction, commerce, and service sectors. The work activities were implemented as a sub-layer where participation is exclusive, i.e., an agent can belong to only one sub-layer (Table 2, the third column). Agents in these sub-layers are aged between 19 and 69 years old and are assigned using a normal distribution. The data used for the creation of this layer are from 2018 edition of the Annual List of Social Information - RAIS [31].

For the creation of the Home layer, the distribution of the Brazilian family size was calculated based on data from Continuous National Household Sample Survey - Continuous PNAD for the fourth quarter of 2019 and the first quarter of 2020 [23], followed by the estimation of the household size probability (Table 3). The distribution was then used to assign each agent to a household of 2 to 13 people.

**Table 3.** Household size and probability distribution for Natal city home layer.

| *Household size* | *Probability* |
| --- | --- |
| 2 | 31.122 |
| 3 | 31.392 |
| 4 | 22.652 |
| 5 | 9.742 |
| 6 | 3.282 |
| 7 | 1.112 |
| 8 | 0.412 |
| 9 | 0.172 |
| 10 | 0.062 |
| 11 | 0.022 |
| 12 | 0.01 |
| 13 | 0.02 |

To model the transport layer, we consider only the public transportation, since it is one of the most populated environments, in which each vehicle has a passenger capacity of 70 people [30]. To create this layer, we used data from the 2010 Brazilian Demographic Census [32], which presents data on the time spent on the journey from home to work or school. Here we used the same methodology as (Ipea, 2013) [41, 42].

Churches are another high-risk environment for the spread of viruses. Here we used two layers for Natal’s most common religions: Catholic and Evangelical. We consider a maximum capacity of 100 people between 0 and 69 years old with a single 2 hours gathering per week. We assigned both layers using a normal distribution. The data for this layer was obtained from the 2010 Brazilian Demographic Census [32].

A separate layer regarding higher education refers to the Federal University of Rio Grande do Norte (UFRN) because it has the largest number of students and workers in the City of Natal (groups of individuals from 12-180). This layer has agents from almost every age range (10-69 years old) and approximately 4.1% of Natal’s population has some connection with UFRN. We used a normal distribution to assign agents to this layer, and the data used comes from the Higher Education Census [34].

The Random layer was implemented to represent any other direct contact between agents, such as drug stores, markets, public parks, etc; or indirect contact, such through objects, surfaces, etc. In this layer, agents are randomly connected regardless of the age range.

#### Decrees

The government’s decrees were the major tool of action of local government to modulate the pandemic course. Decrees included the closure of specific economic and social parts of the society; and the obligatory use of masks. On the model, we implemented the decrees as a change in the probability of contamination (*P*_*contamination*_) and the level of agent interactions (*P*_*interaction*_) that are specific to layers and sub-layers and a specific period. Although all government decrees have their epidemiologic importance, we implemented only the most impactful model adjustment (Table 4). For example, the decree on day March 25th named “Alecrim closure” was an important factor due to this neighbourhood being the major commerce of Natal city. We did not set the *P*_*contamination*_ value to zero because some establishments continued to work.

**Table 4.**
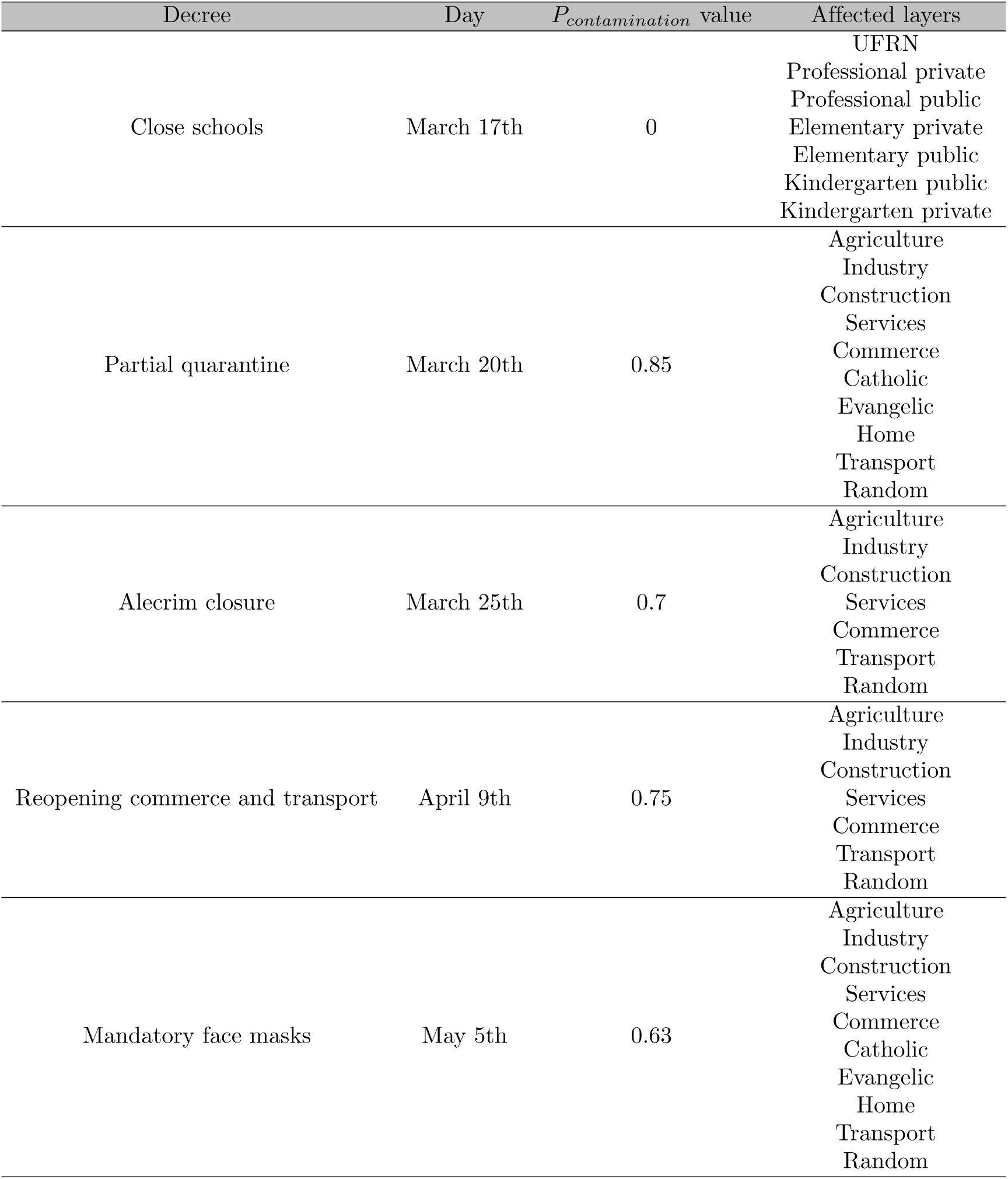
Decrees affect simulation dynamics with variable impact on the different layers and sub-layers over the first wave pandemic. The first column represents the decree’s name. The second column is the implementation day. The third shows the *P*_*contamination*_ value adjusted by the decree. The last column displays the layers and sub-layers affected by the decree.

#### Estimation of external Covid-19 cases and ICU beds

The pandemic starts with initial infections of local individuals by external agents. In order to estimate the number of daily new infections on the first wave pandemic in Natal city due to external interactions, we used daily information about flights and buses, estimated highway traffic and daily reports of confirmed cases.

Natal has only one bus station with an estimated daily number of passengers around 2500 [43]. The pandemic and the decree of March 20, 2020, impacted the bus flow with a decrease by 50% [44]. Until the final of December of 2020, the number of passengers was slowly increasing to around 90% of the regular flow [45]. Thus, we estimated that the total number of bus station passengers during the first wave was 367875.

The number of flights in Natal airport followed the Brazilian pattern. After the restriction measures established in March 2020, an agreement was reached between the aviation companies and ANAC (National Civil Aviation Agency) [46], with a minimum number of flights between the capitals being defined due to the economic infeasibility of maintaining flights with a reduced number of passengers. With the sanitary and economic measures adopted by the government, the number of flights has been slowly returning to normal, reaching 70% of the usual number of flights in December 2020 [47]. Therefore, the number of estimated passengers during the pandemic first wave was around 452233.

Natal is the capital of the state and a metropolitan city, and although we do not have access to the highway flow during the first wave, this number was estimated at 50% of the total cases previously calculated. According to (SESAP-RN), the sum of Covid-19 confirmed cases in the first wave was 26371 (Fig 3A, red line). We used the data to mould the estimated daily external infections (Fig 3A, blue line). First, the daily percentage of infection was obtained by dividing the daily confirmed infections cases by the number of the Natal city population. After that, this percentage was applied to the daily sum of passengers from the bus station and flights, as shown in equation 2.

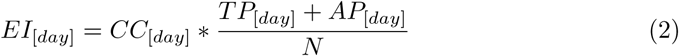

**Fig 3.**
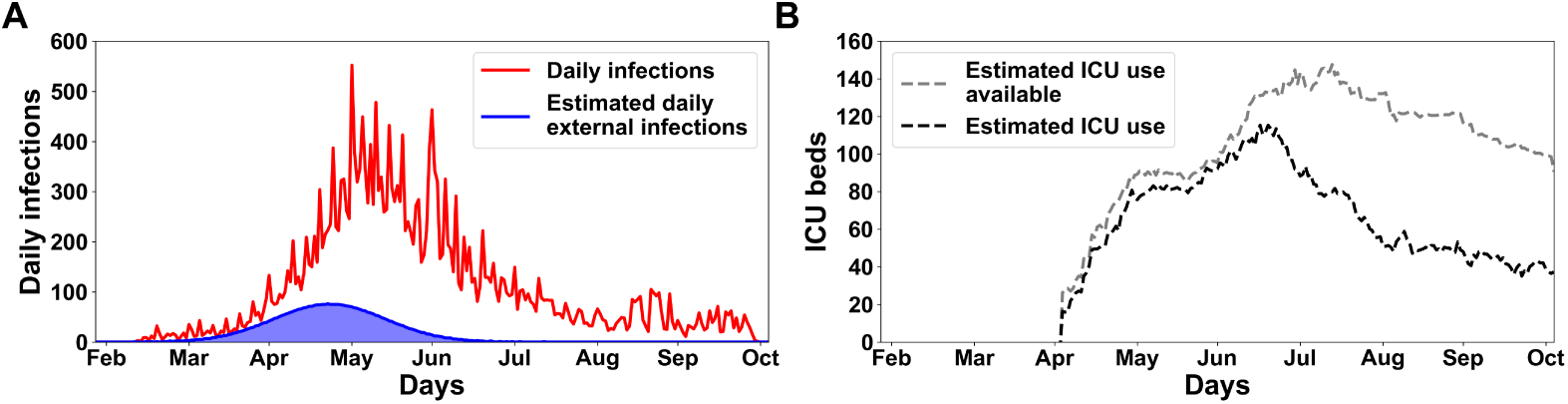
Epidemiological data on the first wave of Covid-19 pandemic in the City of Natal, Brazil. The A panel shows the daily confirmed cases with a total of 26371 and a peak of 552 (red line); and the estimated daily external cases injected into the model with a total of 3957 cases and a peak of 76 cases (blue line). The B panel shows the estimated ICU beds available (silver dashed line) and utilized for the metropolitan region during the first wave (black dashed line), this proportion is 46.68% of real data.

Where *EI*_[*day*]_ is the daily external infections, *CC*_[*day*]_ is the confirmed cases by day, *N* is the total population of Natal, *TP*_[*day*]_ is the daily terrestrial passengers and *AP*_[*day*]_ is the daily air passengers. Thus, the equation 3 calculated the total external cases injected into the model (*TEI*) by adding the estimated number of external infected through highway flow (HF) and the sum of *EI*_[*day*]_ calculated in equation 2. In addition, this external injected cases (total of 3957) were modeled as a gaussian distribution, following the first real infected confirmed case date on March 12th 2020 (Fig 3A, blue line). We opt for not utilized the real number of confirmed cases due to the large absence of tests capable of detecting the Covid-19 pathology in Natal city.

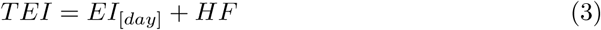

To estimate the ICU bed availability and occupancy for the City of Natal during the first wave of the pandemic, we considered the population size ratio between the metropolitan region and the Rio Grande do Norte state (RN). This metropolitan region is composed of 14 cities, such as: Arês, Ceará-Mirim, Extremoz, Goianinha, Ielmo Marinho, Macaíba, Maxaranguape, Monte Alegre, Natal, Nísia Floresta, Parnamirim, São Gonçalo do Amarante, São José de Mipibu and Vera Cruz. The entire RN state has a population of approximately 3419010 people, and the metropolitan region has around 1596103 inhabitants according to [30], thus, the metropolitan population proportion is 46.68%. This proportion was applied to the actual data acquired from the website (RegulaRN system), which monitors the number of ICU beds available and occupied over the days; for the RN state. The real data were decreased proportionally (46.68%) with respect to the metropolitan region population (Fig 3B).

### Estimation of baseline scenario

In order to fit the simulation outcomes to actual data, were utilized two significant points: (1) The value of *p*_*contamination*_ of each layer/sub-layer and (2) the government decrees. Each decree affects the *p*_*contamination*_ value (increasing or decreasing the value) of different layers/sub-layers in distinct days; consequently, changing the course of the pandemic. Therefore, these points were explored together to reproduce the confirmed daily deaths and deaths accumulated in Natal city during the first wave of the pandemic.

## Results

The first confirmed case of SARS-CoV-2 in the City of Natal was on March 12th 2020 [48]. The number of cases increased and peaked on June 1st 2020, with a 7-day average of 552 new cases per day. The first death was reported on March 31st 2020 [49]. Fatalities follow the growth of new cases to a peak on June 1st 2020, with a 7-day average of 16 deaths per day. The death rate reduced to a 7-day average of new deaths below one event per day in September 2020, returning to an increasing trend in early November 2020. From there on, the first day with no reported deaths by SARS-CoV-2 was on October 2021. This analysis only considers the first wave (initializing on February 26th, two weeks before the first confirmed case, and ending on November 4th). During this wave, the Natal city Secretary for Public Health reported 1072 deaths and 26371 confirmed cases. We focused on the first wave as governmental action was consistently reduced on the second wave that took effect in early 2021, as indicated by the lack of new specific regulatory actions. Moreover, it is impossible to draw a straight comparison between the waves as the second wave was caused by a different virus variant (P1/Gamma) [50], and the vaccine roll-out was concurrent with the second surge [51].

We built an agent-based model to simulate the pandemic evolution through the entire City of Natal with a total of 873383 agents to understand better the impact of governmental interventions on the pandemic. Multiple complex networks connecting different agents were put in place to emulate different layers of social interaction, including schools, work, religious gatherings, transport, commerce and households. Infections due to external agents were estimated from air, road traffic and confirmed infections curve (see Methods). The governmental decrees were implemented as a time-bounded reduction of the specific level of contagiousness. The overall level of contagiousness and a specific factor for each social layer are the open variables subject to modification in the optimisation process. The baseline model uses a set of parameters that provides the most accurate reproduction of the accumulated curve of deaths.

To evaluate the impact of individual decrees, we explored different hypothetical scenarios, including (a) absent interventions, where some decrees have not been applied; (b) effective implementation of the decrees, where the affected layers/sub-layers were fully locked; and (c) delayed interventions, where there were retardment to implement the decrees. We considered three main classes of interventions focused on (1) schools layers, (2) workplaces layers and (3) religious layers. We computed the expected number of deaths for each scenario and compared it with the observed number of deaths in the baseline scenario. For a baseline, absent and effective scenarios, we run a total of 500 simulations. For the delayed intervention scenario, we run a total of 250 simulations for each delay step. We also consider a non-intervention scenario where all decrees were absent with a total of 500 runs. Results are displayed as the mean and standard deviation values for each category unless explicitly noted.

To validate the model, we simulated the first pandemic wave in Natal city in a baseline scenario following the decrees applied (detailed in the previous section and displayed in the figures by the vertical dashed lines). The baseline model could reproduce adequately the daily and accumulated curve of deaths in the first wave (Fig 4A and B, respectively). In the initial months (Feb to early Jun), the simulation outcome followed the curve of deaths actual data. After June, the model results follow closely below the actual data until the start of October, finishing the days simulated with an average of five deaths below the actual data. In total, the observed accumulated number of deaths was 1072, while the number of deaths on the simulation was 1073±52 (Fig 4B). The simulations indicate that a total of 88554 citizens were infected during the first wave, suggesting a sub-notification of about 70.22%. With the model in place, we could inquire about the importance of the different layers for disease propagation by evaluating the amount of new infectious in each social network.

**Fig 4.**
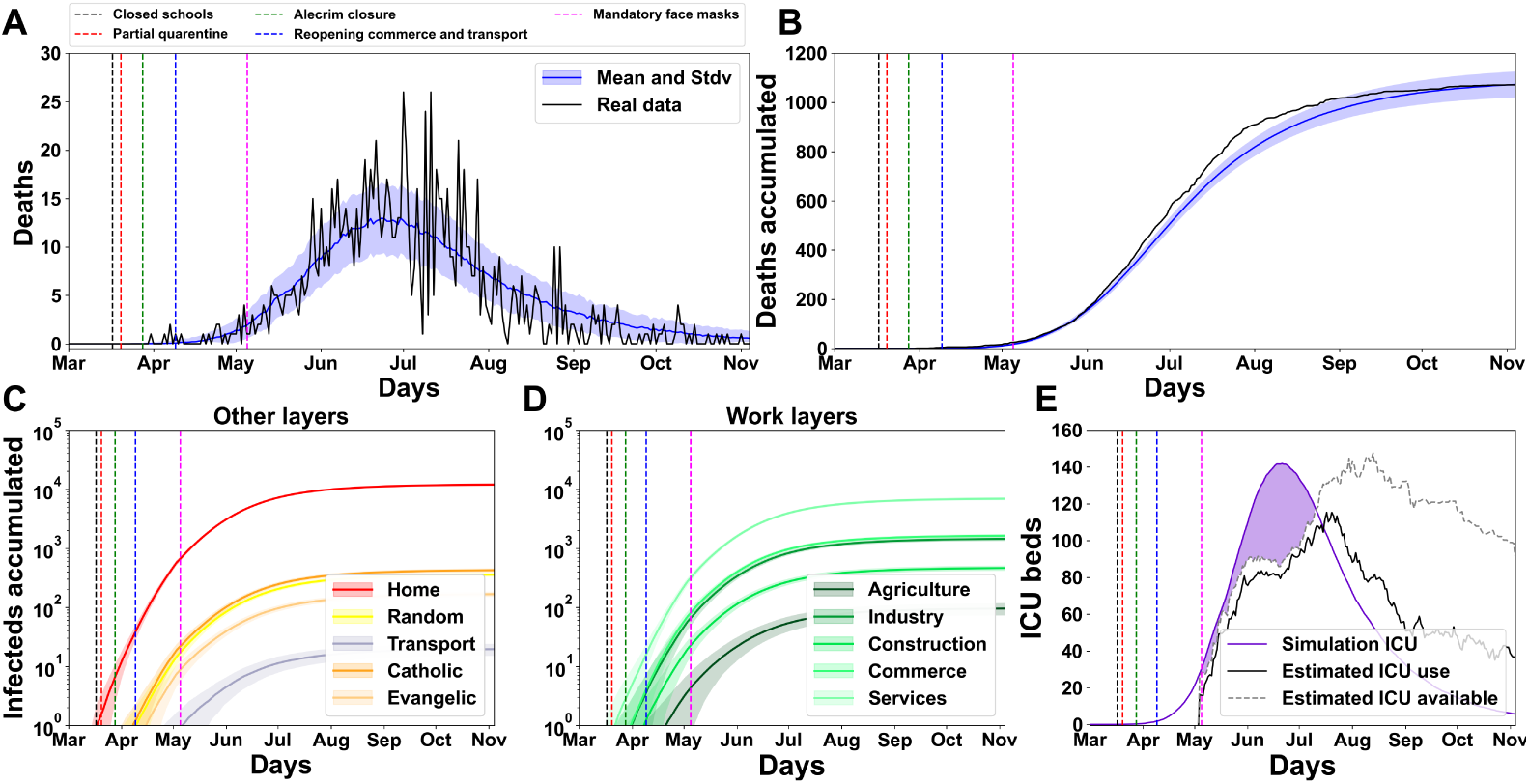
Simulation from the agent model with baseline parameters provides a good fit to epidemiological data on the first wave of SARS-CoV-2 epidemic in the City of Natal. (A) Daily and (B) accumulated deaths during the first wave of SARS-CoV-2 epidemic in the City of Natal (end of February to beginning of October). Real (black line) and simulated data (blue line) are shown. Vertical lines indicate when the governmental decrees became effective. (C and D) The accumulated number of infections in each of the different social layers and sub-layers computed from the model simulation. (E) The estimated daily availability (silver dashed line) and occupation (black line) of ICU beds for the City of Natal. Model prediction of ICU use (solid purple line). Simulated ICU surplus over availability (purple area) indicate that the City of Natal required ICU beds allocated to other municipalities on the state.

The model can quantify the number of infected individuals by layers and sub-layers (Fig4C (Other layers) and D (Work layers)). The most contagious layer was Home (11998 infections) followed by Services (6899) in our simulations. Due to Decree on March 17th 2020, the schools were closed (vertical black dashed line), so there was no infection in these layers. We could also estimate the use of ICU beds by the simulation (Fig4E, purple line) and compare it to the estimated number of ICU beds available for Natal (silver dashed line) and the estimated number of ICU beds use (black line) during the first wave (actual data start on May 4th). In the simulation, the number of ICU beds is unlimited. Therefore, all critical cases that needed ICU were treated. The purple background represents the number of ICUs utilized in the simulation that surpassed the Natal estimated available ICUs (total 7083 and peak of 141 on June 18th). This exceeded ICU occupancy by the simulation can be conjectured by the external arrival of patients searching for a better health system in the state capital.

### Schools layers simulation scenarios

We explored the simulation outcomes with different interventions in the Schools layers and sub-layers (Fig 5). At first, we evaluated the scenario with the absence of the March 17th decree, where the education layers of Natal City were closed (UFRN included), and the maintenance of all other decrees (partial quarantine, Alecrim closure, reopening commerce and transport; and mandatory wearing of face masks) (Fig 5A). There was a considerable increase of daily and accumulated deaths compared with the baseline scenario (Fig 5A, blue line vs black line). Although the initial months (February, March and early April) have similar numbers of death, the following months show a remarkable increase of deaths. Natal would have 6342±199 cumulative deaths, an increase of 5270 when compared with the real outcomes (black line). Natal City reached the highest daily deaths value on July 1st, with 26 deaths reported in the actual data. In this present scenario, the simulated daily deaths reaches its peak on August 13th with an average of 50 daily deaths.

**Fig 5.**
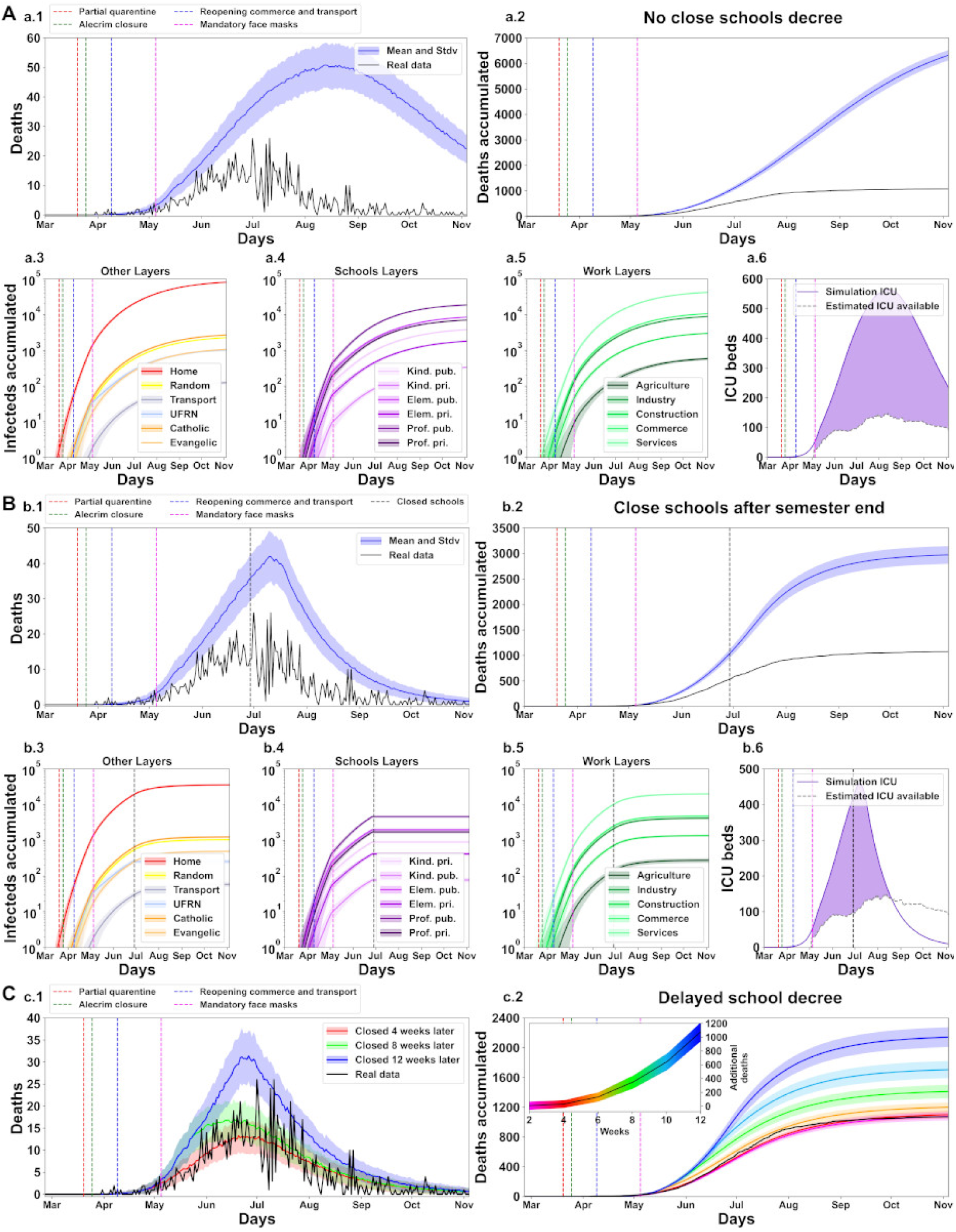
School layers simulation scenarios showing the significant impact of the School decree on March 17th of 2020. Panel description in supporting information 0.1.

By inspecting the evolution of the internal variables, we could assess the number of infections that were due to each of the different layers and sub-layers. The Home layer was the most contaminant (80078 infections) in this scenario, followed by the Services layer (41859). In the Schools layers group, the most contagious sub-layer was Professional public (18545), followed by Elementary public (8457). The most infectious private education sub-layer was Professional private (6979) follow by Kindergarten private (3766). Elementary private (1785), UFRN (1020) and Kindergarten public (332) were less affected. These simulation results suggest that, with the absence of the decree of March 17th, public education would have been the most impacted with infections. All sub-layers of work have an increase in number of infections. Sub-panel a.6 shows the occupancy of ICU beds by the simulations (line and background purple) that overcrowded the estimated number of ICU beds available for Natal (silver dashed line). In this scenario, the occupancy of ICUs by the simulation exceeded the number of ICUs available already on the first day (actual data start on May 4th). This pattern is observed until the end of the simulation with total occupancy of 71485 and reaches its peak on August 11th with 564.

Next, we explored the scenario where the decree of March 17th was postponed and applied on the last day of the school semester on June 29th 2020 (Fig 5B, vertical black dashed line). In this scenario, all Schools layers (UFRN include) would operate normally until the decree date. Besides this, all basic decrees were applied as shown by the vertical dashed lines. The peak of the daily deaths (41) occurs on July 9th (b.1).At the simulations, upon implementing the schools’ closings decree, it takes 12 days (July 11th, one day after the peak) until the decree causes impact and the daily death begins to decrease, however, the simulated daily death continues above the real data line. This decree determines a considerable decrease in cumulative deaths (total of 2970±173, b.2) compared with the a.2 sub-panel for the previous scenario, however, it is still an elevated value when compared with the real data (black line).

In the present scenario, Home (35639 infections) and Services (19858) layers were the most infected. Professional public (4595) and Elementary public (1981) are the most contagious among the schools layers. The Professional private (1706) is the third most infectious and the UFRN occupies fifth place (261). This disposition of the sub-layers most affected is equal to the previous scenario, suggesting that the proposed model is stable. Furthermore, it implies that public education would be the most impacted by the postponed decree. The ICU beds available for Natal City are occupied on the initial day (May 4th) and remains overcrowded until August 20th (peak of 458 at day July 7th and total of 29207), when the occupancy of ICUs by simulation decreases abruptly, showing the impact of the postponed decree. After this day, the use of ICUs was lowered and never exceeded the available number of ICUs again (b.6).

Additionally, we explore the scenarios where the decree of March 17th to close the schools was postponed and applied over different intervals (total of 6) (Fig 5C). Sub-panels c.1 and c.2 show the real data daily accumulated deaths (black line) and the means and standard deviations deaths outcomes of the decree application (colored lines). The c.2 inside graph shows the accumulated curve of additional deaths along the weeks. Postponing this decree would cause an increase of 28±51 additional deaths already in the first four weeks (red line, c.2). This number would abruptly increase to 633±115 in 10 weeks (cyan line, c.2) and 1069±133 in 12 weeks (blue line, c.2).

These simulated scenario outcomes suggest that the closure of the Schools layers group has a major impact over the pandemic course, and that the interval window to government intervention is crucial and narrow; and the decree was applied at an appropriate time.

### Workplaces layers simulation scenarios

In this sub-section, the decrees interventions related to Work layers (Fig 6) were explored. Although the Schools layer has workers, it was decided to consider it only as a School layer, thus, these scenario interventions do not apply to these layers. The first panel illustrates the scenario with the absence of the decree named “Alecrim closure” on March 25th, where the main commercial neighborhood Natal city continues to operate during the first wave of the pandemic [52]. In this scenario, the other decrees (Closed schools, Partial quarantine and Mandatory face masks) were applied, conforming to the real occurrence. Into this scenario would be an increase of deaths accumulated (1235±66) as displayed in a.2 sub-panel, besides this, the number of infections into sub-layers would increase. Among the layers (panel a.3 and a.4), the Home layer was the most impacted with 13879 infection. Services and Commerce sub-layers were the workplaces most affected with 8352 and 1982 infections, respectively. Industry and Construction would finish the first wave with 1749 and 562 and Agriculture would be the work sub-layer less affected with a total of 116 infections. The ICU beds occupancy by the simulation (a.5) overcrowded the available estimative for Natal city (total above 8790); already on the first day (May 4th). In this scenario, the simulation reaches the peak occupancy (total of 157) on June 19th. It maintains this excess until the day June 16th, when decreases below the estimative ICU available for Natal and maintains below until the end of simulation on November 4th.

**Fig 6.**
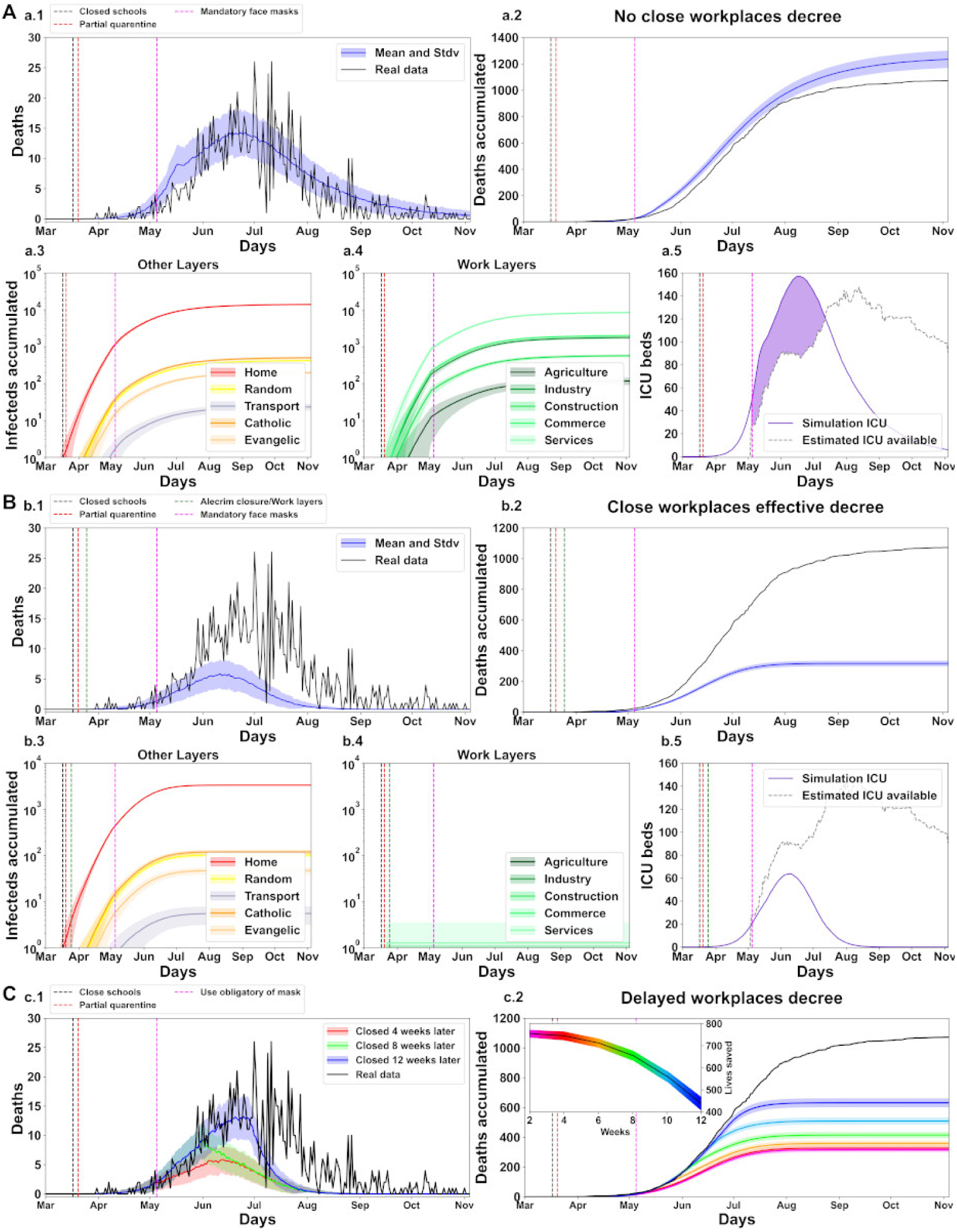
Work layers simulation scenarios showing the impact of the decree on March 25th of 2020. This decree named ”Alecrim closure” defines the closure of the Natal city main commercial neighborhood. Panel description in supporting information 0.1.

It was explored the scenario where the decree on March 25th would have an effective implementation, i.e., the Work layers group would be closed in fact, with the *P*_*contamination*_ value reduced to zero (Fig 6B). The decrees Closed schools, Partial quarantine and Mandatory face masks were applied as shown by the vertical dashed colored lines. In this scenario, the daily death peak occurs on June 14th with a value inferior to 7 deaths (b.1 sub-panel). The deaths accumulated (b.2) were 316±17. Among the layers (sub-panel b.3 and b.4), the Home layer has a decrease of infection with 3335. Although the *P*_*contamination*_ value was reduced to zero into work layers, this decree only applied 16 days after the first confirmed case in Natal city, thus, this was not able to avoid all the infections. Some sub-layers were affected with an average number of infections close to 1 (sub-panel b.3). The absence of infections was an important factor to reduce the occupancy of ICU beds by the simulations. In this scenario, the ICUs peak was on June 6th (total of 63) and did not reach the Natal city estimated ICU use available (sub-panel b.5).

The last panel (Fig 6C) illustrates the scenario of daily (c.1) and cumulative deaths (c.2) where the decree on March 25th would have an effective implementation over different intervals (total of 6). The c.2 inside plot represents the lives saved by the decree over the weeks. The first intervention interval with two weeks did not show a high increase of accumulated deaths (755±16 lives saved). However, the fourth interval (8 weeks, green plot) already presents a visual increase in the cumulative number of deaths and, consequently, a minor number of saved lives (total of 657±21). After 12 weeks (blue plot), the number of lives saved decrease to only 440±29.

These results suggest that with an appropriate decree where the work activities closed, the number of lives saved would be larger and the time to government intervention would be more flexible. In addition, the work layers do not imply a substantial increase in deaths (as the Schools layers). Although, there is a growth in the number of contamination and the ICU beds occupancy when compared with the baseline scenario (Fig 4). This can be conjecture by two points (1) several employees started to work from home (home office), thus, the number of infections decrease in these work social interactions. And (2) for this study was not considered the employees in the informal jobs, these workers possibly were more exposed to the virus and numerically represent an expressive part of the economic sectors as commerce.

### Religious layers simulation scenarios

Religion is an important point in Brazilian culture, in this result sub-section, we explored scenarios with the absence and effective interventions in these sub-layers. The first scenario explored was whether these religious layers (catholic and evangelic) were never been closed (Fig 7A). In the other scenarios, these layers were affected in the decrees named ”Partial Quarantine” and ”Mandatory face masks” where the *P*_*contamination*_ value decreased (Supplementary material), here in this sub-section, the only decree that affects this *P*_*contamination*_ is ”Mandatory face masks”. The decree absence shows a brief increase in the number of daily deaths (a.1) the accumulated deaths (total of 1086±52 a.2). Among the layers (a.3 and a.4 sub-panels), Home has an increase of 162 infections when compared with baseline scenario (Fig 4) and the religious layers catholic and evangelic have an infections increase of 26 (total of 452) and 11 (total of 178), respectively. The a.5 sub-panel shows a minor increase in the occupancy of ICU by the simulation when compared with baseline scenario (Fig 4). This growth was 100 in ICU total occupancy (total above was 7183). This overcrowded reaches its peak on June 18th with 142 and continues the overcrowded until July 10th.

**Fig 7.**
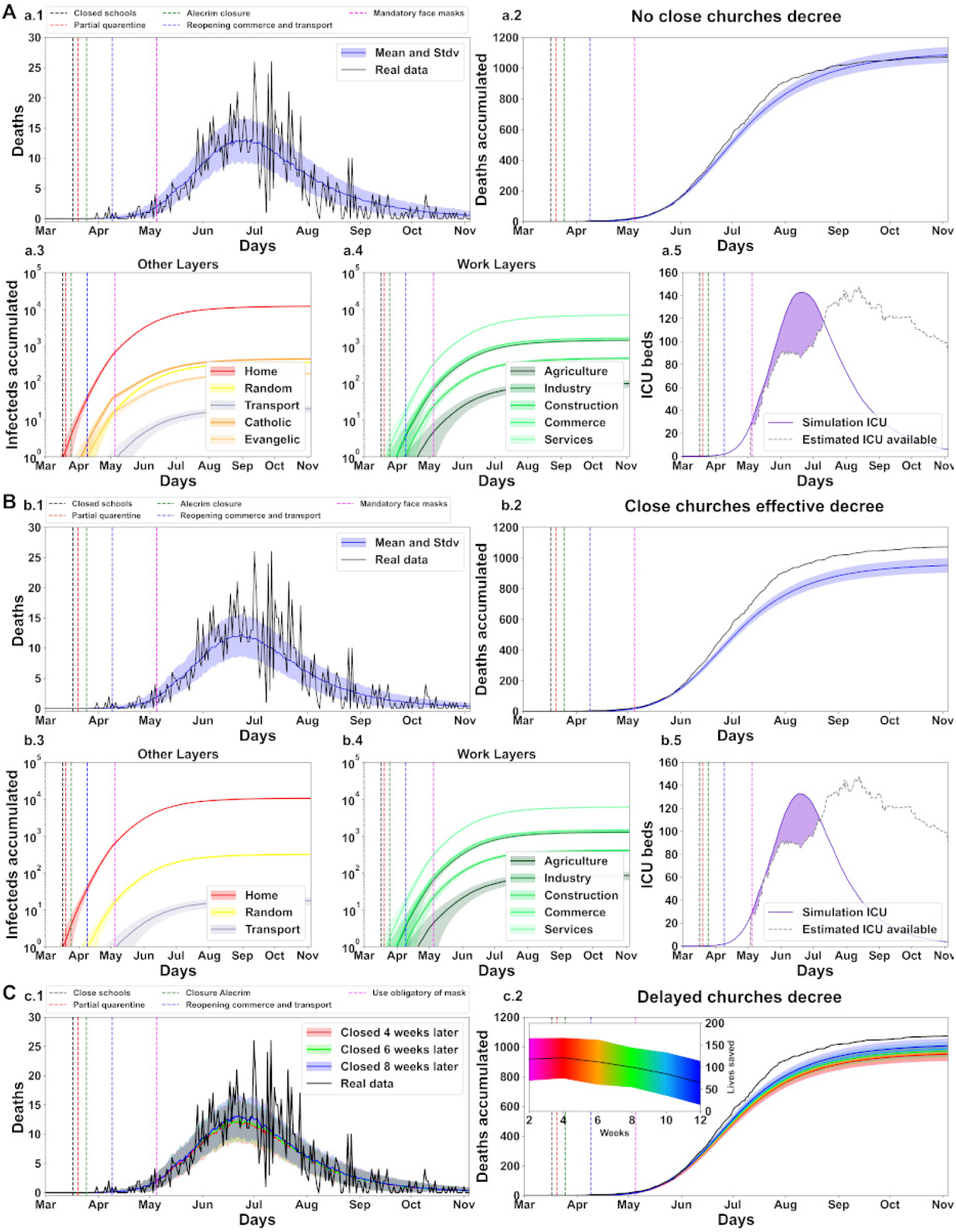
Churches layers simulation scenarios illustrating the impact on the pandemic course.Panel description in supporting information 0.1.

We next explore the scenario where the religious layers were effectively closed by the decree Partial quarantine on the day March 20th (Fig 7B). Sub-panels b.1 and b.2 show the deaths and accumulated deaths (total of 952±47) are inferior to the real data when the religious layers are closed. Among the layers (sub-panels b.3 and b.4), Home has an infection decrease of 1408 (total of 10590). In this scenario, the decree was applied before the disease spread into the religious layers, thus, there were no infections into these layers. Although the closed of religious layers, this intervention was insufficient to prevent the overcrowded ICU beds as shown in b.5 sub-panel. The ICUs occupancy reaches the peak on day June 16th with 132 occupancies.

Finally, we explore the scenarios where the religious layers were authentically closed at different intervals (total of 6) (Fig 7C). The first two intervals (2 and 4 weeks) have do not show a high difference in the number of lives saved, 118±47 and 121±45 respectively. However, this number begins to change on the third intervention interval (6 weeks, 112±50 lives saved) and decreases until the last interval (65±49 lives saved on 12 weeks). These outcomes scenarios suggest that the closure of religious layers decree has a minor impact compared with the previous interventions. This can be assigned to the time brief agents interact in these layers (only two hours per week).

### No-intervention scenario

This subsection illustrates the scenario where the government did not intervene and the Work, Schools and Churches activities were not closed (Fig 8). However, understanding that the critical growing number of cases would alert the population to be careful and even in this scenario would have a constant stimulus to wear masks; we decided to maintain the decree ”Mandatory face masks” on May 5th. The simulations show that, it would have massive daily deaths with a fast increase from April to July where the simulations reaches the peak on July 8th (average of 73 daily death) (Fig 8A). After this month, there is a slow decrease in the following months. The accumulated deaths (total 8000±381) would have an enormous increase of 6927 deaths when compared with the baseline scenario (Fig 4).

**Fig 8.**
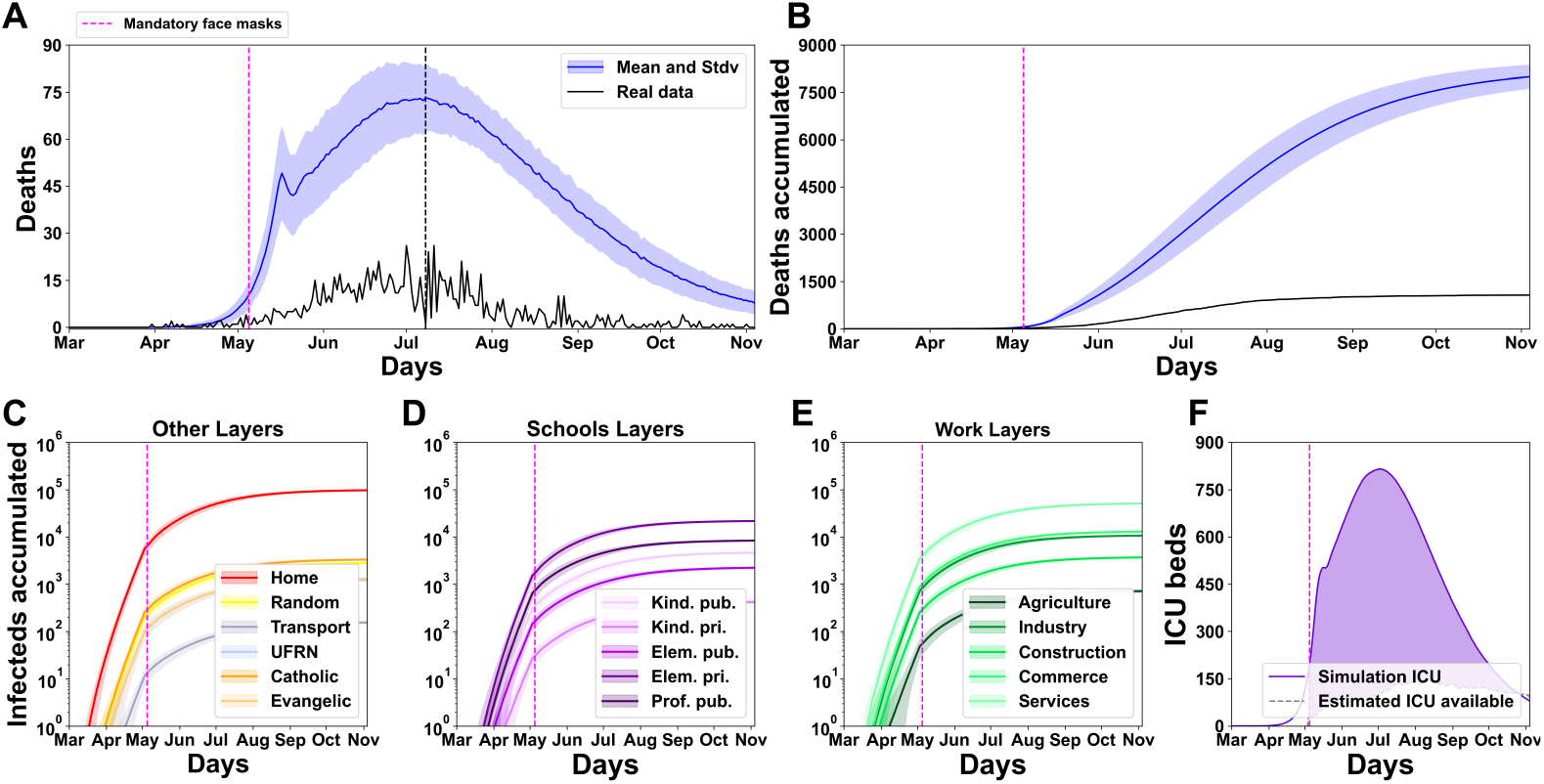
No-intervention scenario showing a catastrophic outcome. Panel description in supporting information 0.1.

All layers have a high increase of infections (Fig 8C, D and E), with Home the most impacted with an total of 97477 infections. Among the Schools layers, Professional public and Elementary public sub-layers were the most contagious and when compared with the scenario with the absence of the closure schools decree (Fig 5A); has an large infections increase of 3212 and 1881 respectively. In the Work layers, Services sub-layer has an extensive increase of 42640 when compared with the scenario with the absence of the Alecrim neighborhood closure (Fig 6A); followed by Commerce and Industry with 11003 and 8928 respectively. Catholic and evangelical has a massive increase of 2863 and 1125 infections respectively, when compared with the absence of the closure churches decree (Fig 7A). The ICU beds occupancy by the simulation overcrowded the available estimative already on the first day of real data and maintains this excess until the end of the simulation on November 4th, reaching the peak on July 1th with 815 (Fig 8F). These results show what can happen in a dangerous scenario. Without the appropriate interventions, the number of deaths would be catastrophic (around 0.91% of Natal population), not to mention the numerous Covid-19 sequels still being studied.

## Discussion

This study presented an agent-based epidemiological model designed to investigate the impact of governmental non-pharmaceutical interventions in the first SARS-CoV-2 wave in the City of Natal (February 26th to November 4th of 2020). The model could support simulations with a 1:1 agents/citizens ratio and used detailed data on (1) health data from daily bulletins report epidemiological, (2) demographic data of Natal city, (3) estimative of external cases arising from airports, bus station and highways flow; and (4) government decrees to confront the pandemic. The model health states extended the SIR model with additional states implemented to represent the complexity of the Covid-19 pathology. We implemented a complex network to emulate the contact among agents. The networks subdivide into layers and sub-layers to better characterise the social interactions such as schools, workplaces, churches, transport, and others, resulting in each network having its own specificities. The simulations successfully reproduced the observed curve of deaths and allowed an estimation of the number of infections and hospitalisations (Fig 4).

Altogether, the results of the simulations support that the actions of state and local governments effectively reduced the loss of citizen lives of the first SARS-CoV-2 wave in the City of Natal. Although the outcome was not optimal - as more rigid interventions could have saved more lives - all interventions seem to have had a positive impact. The model can support the conjecture of hypothetical scenarios with different government interventions. We simulated three main scenarios focused on: (1) school layers, (2) workplaces layers, and (3) religious layers. Table 5 summarises the impact of the interactions among the three main layers groups (schools, workplaces and religions) into each scenario simulated. We identified that the work layers operate as a hub of infections. Although the scenario without a decree for closing workspaces (scenario 6A) resulted in a low increase of deaths, the workspace layers can support a highly infectious dynamic when operating combined with another group of layers, even when the strength of the interaction is low. This effect surges in scenarios without the decrees to close schools (Fig 5A) or churches (Fig 7A). When the interaction into these layers is none, the number of deaths decreases (Fig 6B - Close workplaces effective decree). As the main finding of the simulations with the absence of a Schools closing decree, the model shows a high increase of deaths, around 514.65% (Fig 5A). Nevertheless, the principal finding for workplaces and religious layers was the effective implementation of the decrees. With the absence of activities of the work layers, the number of death decreased by 70.52% (Fig 6B). The closure of religious layers would reduce the ratio of the death by 11.19% (Fig 7B) respectively. The absence of intervention would result in a catastrophic scenario of 8000 deaths, which corresponds to around 0.91% of the entire Natal city population (Fig 8). We can use the model to quantify the number of infected by layers/sub-layers, showing that, typically, the agents get infected primarily on the Home layer. Public education was the most contagious among the School layers simulated scenarios. The Service sub-layer is the work activity most infectious. The most affected religion was Catholicism. These observations strongly support the need for active governmental action in the face of a pandemic such as SARS-CoV-2 ranging from a wide diversity of activities and economic areas.

**Table 5.** Summary of simulation results. Labels: − Low interaction; + High interaction; *X* None interaction; *>* Increase; *<* Decrease; *>>* High increase; *<<* High decrease; *>>>* Higher increase.

|  | Scenarios | Layers |  |  | Outcomes |  |  |
| --- | --- | --- | --- | --- | --- | --- | --- |
|  |  | Schools | Work | Religion | Impact | Deaths | Difference of deaths |
| Fig 4 | Baseline | X | - | - |  | 1073 |  |
| Fig 5A | No “close schools” decree | + | - | - | >> | 6589 | 5516 |
| Fig 6A | No “close workplaces” decree | X | + | - | > | 1234 | 161 |
| Fig 6B | Close workplaces effective | X | X | - | << | 316 | -757 |
| Fig 7A | No “close churches” decree | X | - | + | > | 1086 | 13 |
| Fig 7B | Close churches effective | X | - | X | < | 952 | -121 |
| Fig 8 | No-intervention | + | + | + | >>> | 8000 | 6927 |

We did not analyse the second SARS-CoV-2 wave in the City of Natal in this study. The second wave was longer, deadlier, and more complex, resulting in over 2700 deaths by December 2021. We did not consider the second wave because most of the governmental decrees - but the closure of public schools - became ineffective as the population showed a decreased respect for the measures. As illustrative examples, the Brazilian government decided to proceed with the city elections on November 15th. Traditionally, politicians organise public events for their supporters previously to election day, and although the control of authorities to avoid, these events still occurred [53]. Also, travelling resumed at the end of the year for the two major holidays for the Brazilian population: Christmas and Reveillon [54]. Additionally, there were local reports of several clandestine Carnaval events that occurred on February 2021 [55]. Further, vaccines became progressively widely available in Brazil during the second wave, and, likely, the impact of delayed distribution in the development of the epidemy was more determinant than mobility restrictions. The vaccination in Brazil started in January 2021, prioritising health professionals, older people (60+ years), and indigenous [56]. Although Brazil has a comprehensive and free health system that supports everyone, the population immunisation rhythm was slow due to mismanagement in acquiring and distributing vaccines [57]. The model can be adapted to study the effect of vaccination in the pandemic, but we leave it as a possible follow-up.

## Data Availability

Model software is available at: https://github.com/Sly143/NatalCovid/

## Acknowledgements

PHL, LW, CRC, RCM, LMMV, RKRS, PSS, RCM and PAV received funding from Heriott-Watt University (832228-Singularity/COVID-19 Round 2019-20 GCRF-SFC). The funders had no role in study design, data collection and analysis, decision to publish, or preparation of the manuscript. This research was supported by the High Performance Computing Center at UFRN (NPAD/UFRN)

## Supporting information

### 0.1 Extended labels

Figure 5: (A) Simulations with the decree absence. First line represents the daily and accumulated death of real data (black line in a.1 and a.2) and simulations average and standard deviation (blue line); during the first wave of the pandemic. Other decrees are present and displayed by vertical dashed lines. The cumulative deaths in this scenario have an increase of 591.60% (total 6342±199). Sub-panels a.3 (Other layers), a.4 (Schools layers) and a.5 (Work layers) show the number of infected by layers and sub-layers. Home layer was the most impactful among the layers with an increase of 667.43% when compared with the baseline (Fig 4). The number of Natal available ICUs is exceeded already on the first day of real data record (May 4th) and this overcrowded of cases represent an increase of 1009.25 % when compared with the baseline scenario (Fig 4). (B) Scenario where the Schools close decree was postponed to June 29th. The decrease of daily (b.1) and accumulated deaths (b.2) show the impact of the decree application. The number of deaths was 2970±173; a decrease of 46.83% when compared with the a.2 sub-panel from the previous scenario. Although this wide decrease, this value is still 277.05% higher than the real data (1072). The School layers most affected were Professional public and Elementary public with 4595 and 1981 infections, these values represent a decrease of 75,22% and 76.58% respectively (b.4); when compared with the previous scenario (a.4). The total of cases that overcrowded the ICU beds available were 29207, this number represents a decrease of 59.14% when compared with the previous scenario (a.6), however, this is still an elevated value when compared with the Validated scenario (Fig 4) (an increase of 412.35%). (C) Illustrates the scenario where closing schools decree was applied in differents weeks (total of 6). The first three intervals (until 6 weeks) has a minor impact on the accumulated deaths, however, this number increased considerably from the fourth interval (8 weeks), reaching the mark of 1069±133 accumulated deaths in 12 weeks.

Figure 6: (A) Shows the scenario with the absence of the decree with daily (a.1) and accumulated deaths (a.2) increasing 15.21% when compared with baseline. Among the layers (a.3 and a.4), Home has an increase of 15.68% (total of 13879) and was the most impacted. The infections of Work layers group increased an average of 21.4%, with the Service sub-layer the most affected with a total of 8352 (increase of 1453 cases) followed by Commerce with 1982 (increase of 349). The sub-layer less affected was the Agriculture layer with an increase of 21 cases (total of 116 cases). In this scenario, the occupancy number of ICU beds by the simulation increase 24.1%. (B) Presents the scenario with the effective implementation of the decree on day March 25th, where all Work layers were closed. The accumulated deaths (b.2) were 316±17 showing a decrease of 70.52% when compared with the baseline scenario (Fig 4). The layer Home (b.3) was the most infectious with 3335 case (decrease of 72.2%) follow by catholic with 118 (decrease of 72.3%). The decree, although effective, was not able to completely annul the contamination into the work layer group (b.4), in reason the date of implementation. However, the effectiveness was almost perfect with the contamination close to 1 case. The effective implementation of the decree was sufficient to reduce the number of ICUs occupancy below the number of ICUs available (b.5). The peak of occupancy was only 63 (June 6th); on this day, the number of available ICUs was 90 (remain 27 available). (C), illustrates the exploration of the Alecrim closure decree authentic application in different intervals (total of 6). Similar to Schools layers delay decree, over the first three intervals (six weeks) of decree delay, there was a minor increase in accumulated death (c.2). However, this value increases fast into the next three intervals. Thus, fewer lives would be saved as shown in the inside graph of c.2.

Figure 7: (A) Show the scenario with the religious layers (catholic and evangelic) function normally. The deaths accumulated (a.2) increase 1.21%(total of 1086±5) when compared with the baseline scenario (Fig 4) (1073). Among the layers (a.3 and a.4), Home has an increase of 1.35% (total of 12160 infections) and the religious catholic and evangelic grows 6.1% (452) and 6.59% (178) cases, respectively. The ICU overcrowded (a.5) increase 1.41%. (B) Presents the scenario where the religious layers were effectively closed. (b.2) Show a great reduction in the number of deaths accumulated (11.28%) at the end of the first wave (total of 952±47) when compared with the baseline scenario (Fig 4). Sub-panels b.3 and b.4 show the layers infections, Home was the most affected with a reduction of 11.74% (total 10590 cases). The religious catholic and evangelic layers did not have contamination. Even with the decree, the occupancy of ICU beds by the simulation was superior to the available estimated (a.5) with peak of 132 on June 16th. (C) illustrates the scenario where the religious layers closed decree was applied indeed at different intervals (total of 6). All intervals present minor impact over the death accumulative when compare with previous scenarios.

Figure 8: The daily (A) and accumulated deaths (B) are would have a high increase when compare with baseline scenario (Fig 4), the total of accumulated deaths (8000) represents an increase of 646.27%. All layers and sub-layers have a great increase of contamination, Home was the most affected with 97477 infections (an increase of 712.44%) when compared with baseline scenario (Fig 4). Among the work layers, the most infected was Services sub-layer with 50992 (increase of 510.54%) follow by the Commerce sub-layer with 12985 (increase of 555.15%) when compared with the scenario with the absence of the closure work layers decree (Fig 6A). When compared with the scenario with the decree for never closing schools (Fig 5A), Professional public has an increase of 17.32% (total 21757) follow by Elementary public with an increase of 22.24% (10338). The religions catholic and evangelical have an increase of 633.41% (3315) and 632.02% (1303) respectively, when compared with the absence of the closure churches decree scenario (Fig 7A). The peak of ICU occupancy (815) was 478.01% higher than the baseline scenario Fig 4 scenario and the total above the available ICU (88028) would be sufficient to overcrowd the ICUs available throughout the state during the first wave of the SARS-CoV-2.

### 0.2 Supplementary tables

**Table 6.** Decrees utilized in Schools layers simulation Fig 5A - Never close schools sub-layers.

| Decree | Day | P contamination value | Affected layers |
| --- | --- | --- | --- |
| Partial quarantine | March 20th | 0.85 | Agriculture<br>Industry<br>Construction<br>Services<br>Commerce<br>Catholic<br>Evangelic<br>Home<br>Transport<br>Random |
| Alecrim closure | March 25th | 0.7 | Agriculture<br>Industry<br>Construction<br>Services<br>Commerce<br>Transport<br>Random |
| Reopening commerce and transport | April 9th | 0.75 | Agriculture<br>Industry<br>Construction<br>Services<br>Commerce<br>Transport<br>Random |
| Use obligatory of mask | May 5th | 0.63 | Agriculture<br>Industry<br>Construction<br>Services<br>Commerce<br>Catholic<br>Evangelic<br>Home<br>Transport<br>Random<br>Professional private<br>Professional public<br>Elementary private<br>Elementary public<br>Kindergarten public<br>Kindergarten private<br>UFRN |

**Table 7.**
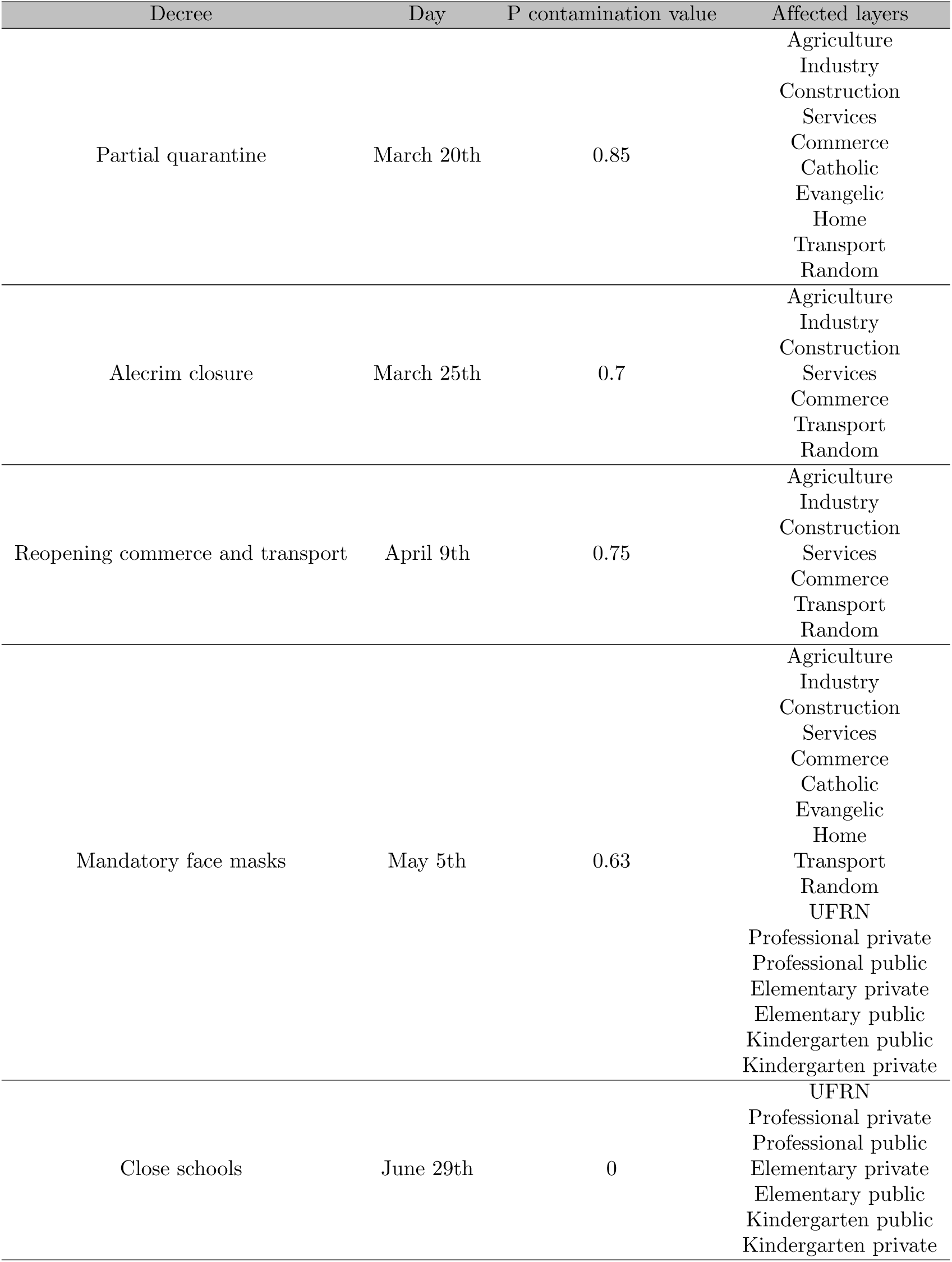
Decrees utilized in Schools layers simulation Fig 5B - Close schools sub-layers after semester ends.

**Table 8.**
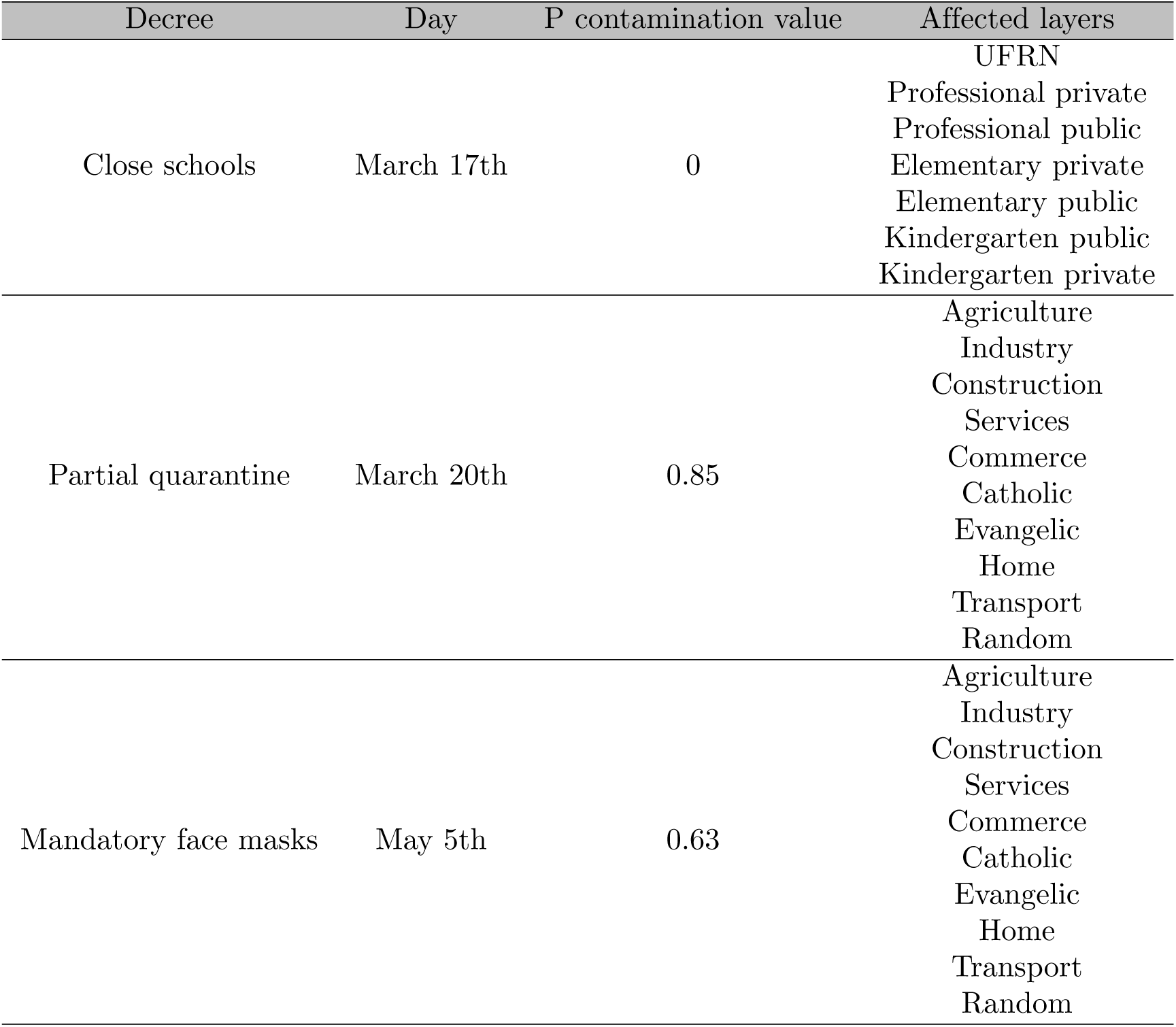
Decrees utilized in Work layers simulation Fig 6A - Never close work sub-layers.

**Table 9.**
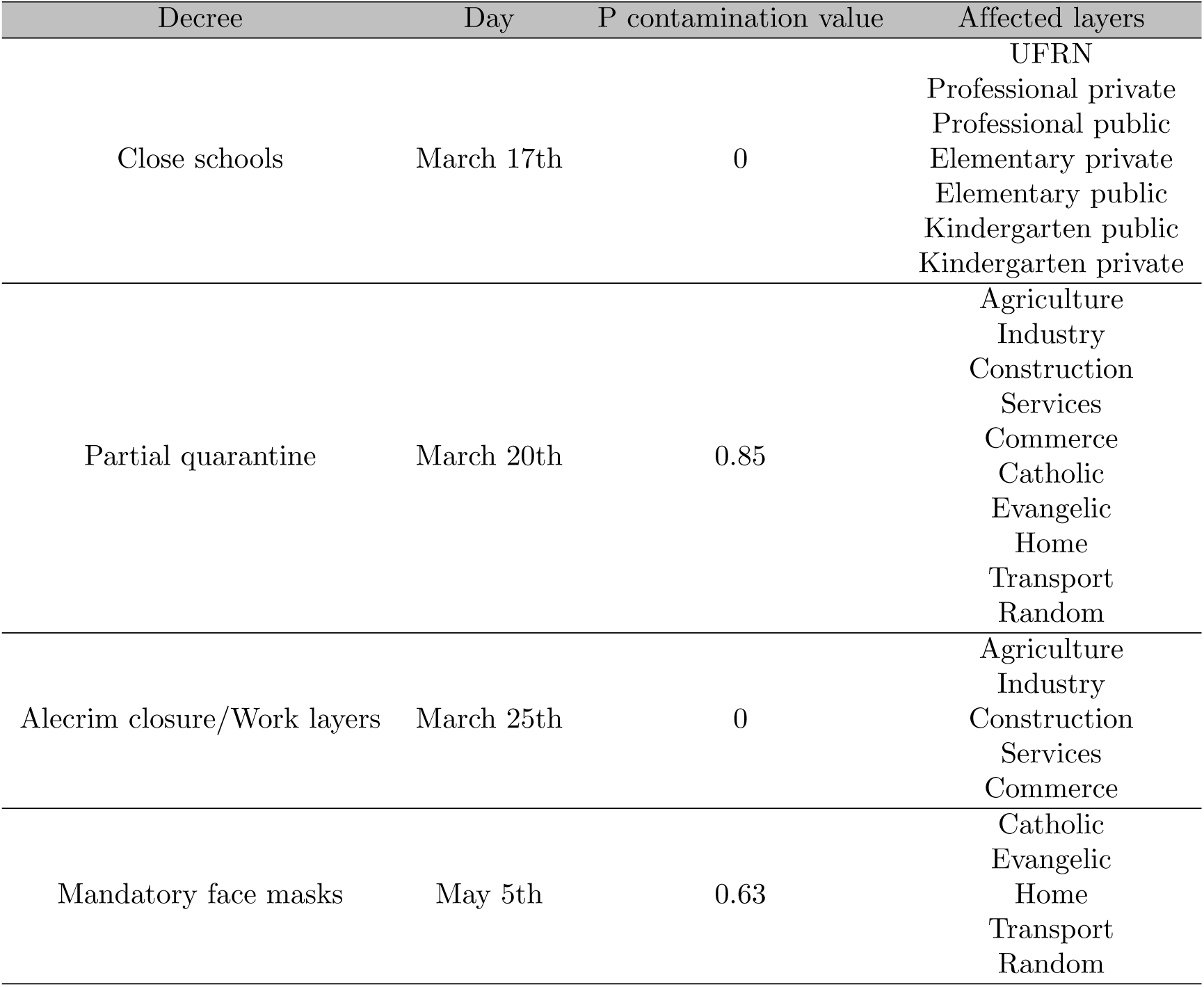
Decrees utilized in Work layers simulation Fig 6B - Really close work sub-layers.

**Table 10.** Decrees utilized in Churches layers simulation Fig 7A - Never close churches sub-layers.

| Decree | Day | P contamination value | Affected layers |
| --- | --- | --- | --- |
| Close schools | March 17th | 0 | UFRN<br>Professional private<br>Professional public<br>Elementary private<br>Elementary public<br>Kindergarten public<br>Kindergarten private |
| Partial quarantine | March 20th | 0.85 | Agriculture<br>Industry<br>Construction<br>Services<br>Commerce<br>Home<br>Transport<br>Random |
| Alecrim closure | March 25th | 0.7 | Agriculture<br>Industry<br>Construction<br>Services<br>Commerce<br>Transport<br>Random |
| Reopening commerce and transport | April 9th | 0.75 | Agriculture<br>Industry<br>Construction<br>Services<br>Commerce<br>Transport<br>Random |
| Mandatory face masks | May 5th | 0.63 | Agriculture<br>Industry<br>Construction<br>Services<br>Commerce<br>Catholic<br>Evangelic<br>Home<br>Transport<br>Random |

**Table 11.**
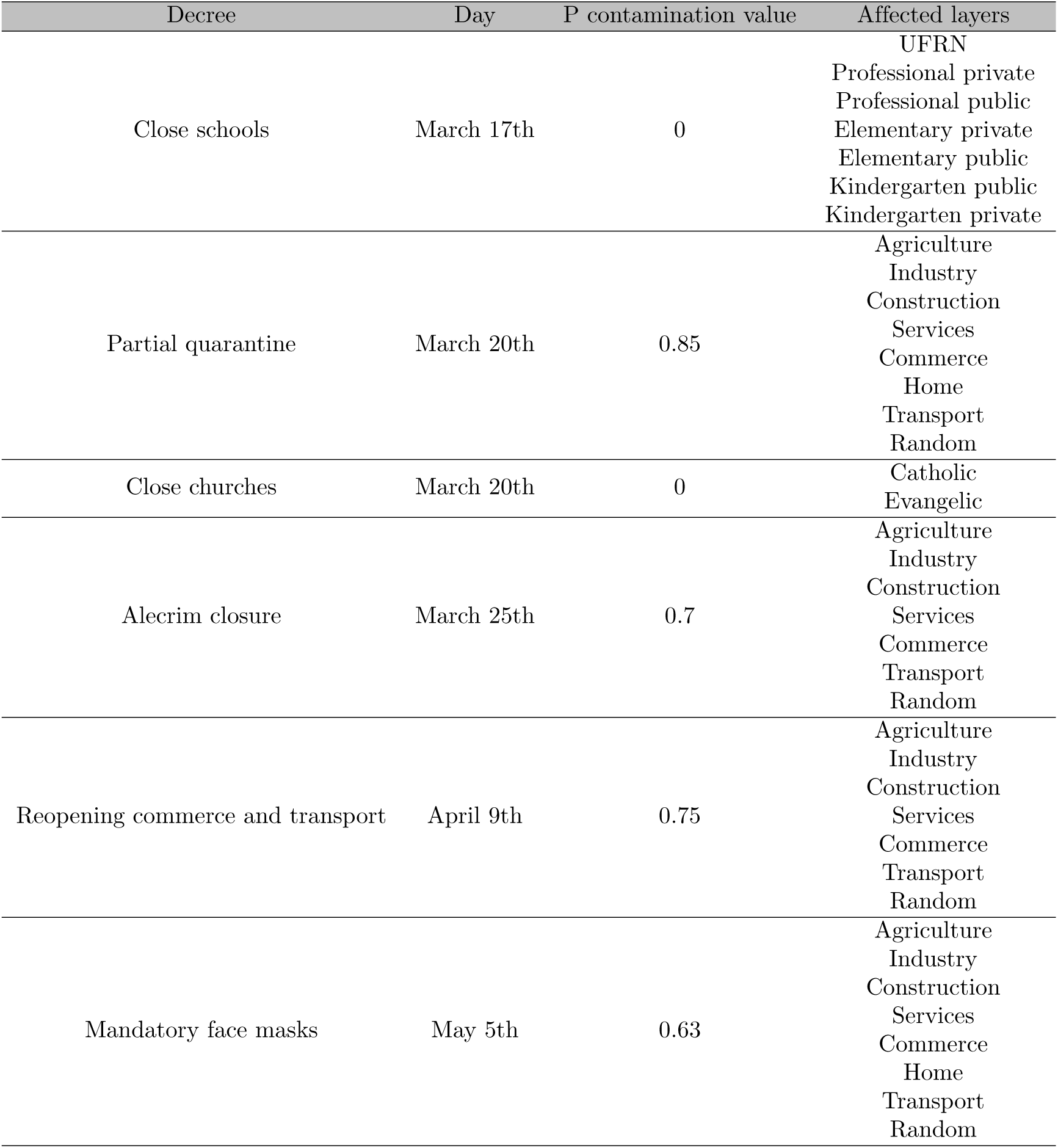
Decrees utilized in Churches layers simulation Fig 7B - Really close churches sub-layers.

## References

1. Wu F, Zhao S, Yu B, Chen YM, Wang W, Song ZG, et al. A new coronavirus associated with human respiratory disease in China. Nature. 2020;579(7798):265–269. doi:10.1038/s41586-020-2008-3.

2. Zhou P, Yang XL, Wang XG, Hu B, Zhang L, Zhang W, et al. A pneumonia outbreak associated with a new coronavirus of probable bat origin. Nature. 2020;579(7798):270–273. doi:10.1038/s41586-020-2012-7.

3. Andersen KG, Rambaut A, Lipkin WI, Holmes EC, Garry RF. The proximal origin of SARS-CoV-2; 2020. Available from: https://doi.org/10.1038/s41591-020-0820-9.

4. Cucinotta D, Vanelli M. WHO declares COVID-19 a pandemic. Acta Biomedica. 2020;91(1):157–160. doi:10.23750/abm.v91i1.9397.

5. Verity R, Okell LC, Dorigatti I, Winskill P, Whittaker C, Imai N, et al. Estimates of the severity of coronavirus disease 2019: a model-based analysis. The Lancet Infectious Diseases. 2020;20(6):669–677. doi:10.1016/S1473-3099(20)30243-7.

6. Wu JT, Leung K, Bushman M, Kishore N, Niehus R, de Salazar PM, et al. Estimating clinical severity of COVID-19 from the transmission dynamics in Wuhan, China. Nature Medicine. 2020;26(4):506–510. doi:10.1038/s41591-020-0822-7.

7. Russell TW, Hellewell J, Jarvis CI, van Zandvoort K, Abbott S, Ratnayake R, et al. Estimating the infection and case fatality ratio for coronavirus disease (COVID-19) using age-adjusted data from the outbreak on the Diamond Princess cruise ship, February 2020. Eurosurveillance. 2020;25(12):2000256. doi:10.2807/1560-7917.ES.2020.25.12.2000256.

8. Wang H, Paulson KR, Pease SA, Watson S, Comfort H, Zheng P, et al. Estimating excess mortality due to the COVID-19 pandemic: a systematic analysis of COVID-19-related mortality, 2020–21. The Lancet. 2022;.

9. Clemens J, Aziz AB, Tadesse BT, Kang S, Marks F, Kim J. Evaluation of protection by COVID-19 vaccines after deployment in low and lower-middle income countries. EClinicalMedicine. 2022;43:101253.

10. Editorial. Why a pioneering plan to distribute COVID vaccines equitably must succeed. Nature. 2021;589(7841):170–170.

11. Heneghan CJ, Spencer EA, Brassey J, Plüddemann A, Onakpoya IJ, Evans DH, et al. SARS-CoV-2 and the role of airborne transmission: a systematic review. F1000Research. 2021;10:232. doi:10.12688/f1000research.52091.1.

12. Greenhalgh T, Jimenez JL, Prather KA, Tufekci Z, Fisman D, Schooley R. Ten scientific reasons in support of airborne transmission of SARS-CoV-2. The Lancet. 2021;397(10285):1603–1605. doi:10.1016/S0140-6736(21)00869-2.

13. Kutter J, de Meulder D, Bestebroer T, et al. SARS-CoV and SARS-CoV-2 are transmitted through the air between ferrets over more than one meter distance. Nat Commun. 2021;12:1–8.

14. Johansson M, Quandelacy T, Kada S, et al. SARS-CoV-2 transmission from people without COVID-19 symptoms. JAMA Netw Open. 2021;4.

15. Instituto Brasileiro de Geografia e Estatística. Estimativas de populąco; 2020. https://www.ibge.gov.br/estatisticas/sociais/populacao/9103-estimativas-de-populacao.html?=&t=o-que-e.

16. Candido DS, Claro IM, de Jesus JG, Souza WM, Moreira FRR, Dellicour S, et al. Evolution and epidemic spread of SARS-CoV-2 in Brazil. Science. 2020;369(6508):1255–1260. doi:10.1126/SCIENCE.ABD2161.

17. Dong E, Du H, Gardner L. An interactive web-based dashboard to track COVID-19 in real time. The Lancet infectious diseases. 2020;20(5):533–534.

18. Instituto Brasileiro de Geografia e Estatística. IBGE Cidades; 2021. https://cidades.ibge.gov.br/.

19. Kraemer MUG, Yang CH, Gutierrez B, Wu CH, Klein B, Pigott DM, et al. The effect of human mobility and control measures on the COVID-19 epidemic in China. Science (New York, NY). 2020;497(May):493–497. doi:10.1126/science.abb4218.

20. Kissler, S, Tedijanto, C, Goldstein, E, Grad, Y, Lipsitch M. Projecting the transmission dynamics of SARS-CoV-2 through the postpandemic period. Science. 2020;5793(April).

21. Ferguson NM, Laydon D, Nedjati-gilani G, Imai N, Ainslie K, Baguelin M, et al. Report 9 : Impact of non-pharmaceutical interventions (NPIs) to reduce COVID-19 mortality and healthcare demand. Imperial College Online Report. 2020;(March).

22. Pnud Brasil, Ipea e FJP. Atlas do Desenvolvimento Humano no Brasil; 2021. http://www.atlasbrasil.org.br/.

23. Instituto Brasileiro de Geografia e Estatística. Continuous National Household Sample Survey - Continuous PNAD; 2020. https://www.ibge.gov.br/estatisticas/sociais/populacao/9173-pesquisa-nacional-por-amostra-de-domicilios-continua-trimestrahtml?edicao=27704&t=destaques.

24. Lyra W, do Nascimento JD, Belkhiria J, de Almeida L, Chrispim PP, de Andrade COVID-19 pandemics modeling with SEIR(+CAQH), social distancing, and age stratification. The effect of vertical confinement and release in Brazil. medRxiv. 2020; p. 2020.04.09.20060053. doi:10.1101/2020.04.09.20060053.

25. Lemos-Paião AP, Silva CJ, Torres DFM. A New Compartmental Epidemiological Model for COVID-19 with a Case Study of Portugal. Ecological Complexity. 2020;44:100885. doi:10.1016/j.ecocom.2020.100885.

26. Leontitsis A, Senok A, Alsheikh-Ali A, Nasser YA, Loney T, Alshamsi A. Seahir: A specialized compartmental model for covid-19. International Journal of Environmental Research and Public Health. 2021;18(5):1–11. doi:10.3390/ijerph18052667.

27. Dashtbali M, Mirzaie M. A compartmental model that predicts the effect of social distancing and vaccination on controlling COVID-19. Scientific Reports. 2021;11(1):8191. doi:10.1038/s41598-021-86873-0.

28. Perez L, Dragicevic S. An agent-based approach for modeling dynamics of contagious disease spread. International Journal of Health Geographics. 2009;8(1):1–17. doi:10.1186/1476-072X-8-50.

29. Hunter E, Namee BM, Kelleher J. An open-data-driven agent-based model to simulate infectious disease outbreaks. vol. 13; 2018.

30. Municipal Department of Environment and Urbanism; 2021. https://www.natal.rn.gov.br/semurb.

31. Ministry of Economy. Annual List of Social Information – RAIS; 2018. http://pdet.mte.gov.br/microdados-rais-e-caged.

32. Instituto Brasileiro de Geografia e Estatística. Brazilian Demographic Census; 2010. https://www.ibge.gov.br/estatisticas/sociais/populacao/9662-censo-demografico-2010.html?=&t=o-que-e.

33. Instituto Nacional de Estudos e Pesquisas Educacionais Anísio Teixeira. Brazilian School Census; 2019. https://www.gov.br/inep/pt-br/acesso-a-informacao/dados-abertos/microdados/censo-escolar.

34. Instituto Nacional de Estudos e Pesquisas Educacionais Anísio Teixeira. Higher Education Census; 2018. https://www.gov.br/inep/pt-br/acesso-a-informacao/dados-abertos/microdados/censo-da-educacao-superior.

35. Kermack WO, McKendrick AG. A Contribution to the Mathematical Theory of Epidemics. Proceedings of the Royal Society of London Series A, Containing Papers of a Mathematical and Physical Character. 1927;115(772):700–721.

36. Scabini LFS, Ribas LC, Neiva MB, Junior AGB, Farfán AJF, Bruno OM. Social interaction layers in complex networks for the dynamical epidemic modeling of COVID-19 in Brazil. Physica A: Statistical Mechanics and its Applications. 2021;564:125498. doi:10.1016/j.physa.2020.125498.

37. Yanes-Lane M, Winters N, Fregonese F, Bastos M, Perlman-Arrow S, Campbell JR, et al. Proportion of asymptomatic infection among COVID-19 positive persons and their transmission potential: A systematic review and meta-analysis. PloS one. 2020;15(11):e0241536.

38. Lauer SA, Grantz KH, Bi Q, Jones FK, Zheng Q, Meredith HR, et al. The incubation period of coronavirus disease 2019 (COVID-19) from publicly reported confirmed cases: estimation and application. Annals of internal medicine. 2020;172(9):577–582.

39. Zayet S, Gendrin V, Klopfenstein T. Natural history of COVID-19: back to basics. New Microbes and New Infections. 2020;38:100815.

40. Kacem I, Gharbi A, Harizi C, Souissi E, Safer M, Nasri A, et al. Characteristics, onset, and evolution of neurological symptoms in patients with COVID-19. Neurological Sciences. 2021;42(1):39–46.

41. Microdados dos Censo Demografico 2010; 2010. https://sidra.ibge.gov.br/tabela/3604.

42. Pereira RHM, Schwanen T. Tempo de deslocamento casa-trabalho no Brasil (1992-2009): diferenças entre regiões metropolitanas, níveis de renda e sexo. Texto para Discussão; 2013.

43. Website of Natal bus station; 2020. https://www.rodoviarianatal.com.br/sobre-o-terminal/.

44. Government RN’s decrees during the pandemic; 2021. https://portalcovid19.saude.rn.gov.br/medidas/medidasdogoverno/.

45. Bus station holiday report; 2020. https://www.rodoviarianatal.com.br/2020/12/22/20-mil-pessoas-devem-viajar-nos-feriados-do-final-do-ano-a-partir-do-terminal-rodoviario-de-natal/

46. Report decrease flights during first wave; 2020. https://www1.folha.uol.com.br/mercado/2020/03/companhias-reduzem-malha-aerea-para-um-voo-diario-para-cada-capital-por-coronavirus.shtml.

47. Report increase flights during first wave; 2020. https://www.gov.br/infraestrutura/pt-br/assuntos/noticias/em-dezembro-brasil-atinge-70-da-quantidade-de-voos-na-comparacao-co

48. First confirmed case; 2020. https://g1.globo.com/rn/rio-grande-do-norte/noticia/2020/03/12/rn-tem-primeiro-caso-confirmado-do-novo-coronavirus-diz-secretaria-secretaria-estadual-de-saude.ghtml.

49. First confirmed death; 2020. https://g1.globo.com/rn/rio-grande-do-norte/noticia/2020/03/31/secretaria-de-saude-confirma-segunda-morte-por-coronavirus-no-rn.ghtml.

50. Nicolete VC, Rodrigues PT, Fernandes AR, Corder RM, Tonini J, Buss LF, et al. Epidemiology of COVID-19 after Emergence of SARS-CoV-2 Gamma Variant, Brazilian Amazon, 2020–2021. Emerging infectious diseases. 2022;28(3):709.

51. Lotta G, Fernandez M, Kuhlmann E, Wenham C. COVID-19 vaccination challenge: what have we learned from the Brazilian process? The Lancet Global Health. 2022;.

52. Closure of Alecrim neighborhood; 2020. https://g1.globo.com/rn/rio-grande-do-norte/noticia/2020/03/25/coronavirus-maior-centro-de-comercio-popular-de-natal-alecrim-amanhence-com-lojas-fechadas-e-ruas-vazias.ghtml.

53. Elections public events; 2020. https://oglobo.globo.com/politica/eleicoes-2020/na-contramao-da-pandemia-candidatos-mantem-campanha-de-rua-1-247364

54. Revellion in Brazil; 2021. https://g1.globo.com/fantastico/noticia/2021/01/03/festas-na-pandemia-especialistas-alertam-sobre-os-efeitos-das-aglomeracoes-do-fim-do-ano.ghtml.

55. Carnaval public events; 2021. https://g1.globo.com/rn/rio-grande-do-norte/noticia/2021/02/14/covid-19-mesmo-com-proibicao-de-festas-de-carnaval-aglomeracoes-registradas-em-natal-e-pipa.ghtml.

56. Vaccination start in Brazil; 2021. https://agenciabrasil.ebc.com.br/saude/noticia/2021-01/vacinacao-contra-covid-19-come%C3%A7a-em-todo-o-pais.

57. Xavier DR, Lima e Silva E, Lara FA, e Silva GRR, Oliveira MF, Gurgel H, et al. Involvement of political and socio-economic factors in the spatial and temporal dynamics of COVID-19 outcomes in Brazil: A population-based study. The Lancet Regional Health â€“ Americas. XXXX;doi:10.1016/j.lana.2022.100221.

